# How varying intervention, vaccination, mutation and ethnic conditions affect COVID-19 resurgence

**DOI:** 10.1101/2021.08.31.21262897

**Authors:** Longbing Cao, Qing Liu

## Abstract

After a year of the unprecedented COVID-19 pandemic in 2020, the world has been overwhelmed by COVID-19 resurgences and virus mutations up to today. Here we develop a dynamic intervention, vaccination and mutation-driven epidemiological model with sequential interventions influencing epidemiological compartments and their state transition. We quantify epidemiological differences between waves under fatal viral mutations, the impacts of control or relaxation interventions and fatal virus mutations on resurgence under vaccinated or unvaccinated conditions, and estimate potential trends under varying interventions and mutations. Comprehensive analyses - between waves, with or without vaccinations, across representative countries with distinct ethnic and cultural backgrounds, what-if scenario simulations on second waves, and future 30-day trend - in two COVID-19 waves in Germany, France, Italy, Israel and Japan over 2020 and 2021 obtain quantitative empirical indication of the influence of strong vs. weak interventions, various combinations of control vs. relaxation strategies, and different transmissibility levels of coronavirus mutants on the behaviors and patterns of different waves and resurgences and future infection trends. The analyses quantify that (1) virus mutations, intervention fatigue, early relaxations, and lagging interventions, etc. may be common reasons for the resurgences observed in many countries; (2) timely strong interventions such as full lockdown will contain resurgence; (3) some resurgences relating to fatal mutants could have been better contained by either carrying forward the effective interventions from their early waves or implementing better controls and timing; (4) insufficient evidence is found on distinguishing the infection between unvaccinated and vaccinated countries while substantial vaccinations ensure much low mortality rate and high recovery rate; (5) resurgences with substantial vaccination have a much lower mortality rate and a higher recovery rate than those without vaccination; and (6) in the absence of sufficient vaccination, herd immunity and effective antiviral pharmaceutical treatments and with more infectious mutations, the widespread early or fast relaxation of interventions including public activity restrictions likely result in a COVID-19 resurgence. We also find the severity, number and timing of control and relaxation interventions determines a protection-deconfinement tradeoff, which can be used to evaluate the containment effect and the opportunity of resurgence and reopening under vaccination and fatal mutations.

---

The COVID-19 pandemic has swept across the world, infecting more than 647 million people and causing over 6.6 million deaths^1^. The very first waves in most jurisdictions were successfully managed by immediately and harshly enforcing various non-pharmaceutical interventions (NPIs, for simplicity, we refer ‘intervention’ to strategies, policies, countermeasures and restrictions to contain the epidemic), such as social distancing, face mask wearing, lockdowns, and travel bans. As early as April 2020, there were warnings against the premature relaxation of strict interventions^1^ and the potential risk of a second wave of infections^2^. Unfortunately, many areas that were successful in containing the first wave have suffered from second and multiple waves and hotspot-based resurgences. The resurgence transform is further strengthened by rapid SARS-CoV-2 mutations such as the delta, lambda, mu and omicron mutants with substantially higher transmissibility and more sophisticated epidemiological attributes. While our understandings of both the SARS-CoV-2 virus and the COVID-19 disease and the influence of interventions are now much deeper and more comprehensive than in 2020^3^, we are short of quantitative evidence about the interactive influence of different severity, number and timing of control versus relaxation interventions with increasingly more infectious mutants on the resurgence of COVID-19, the different behaviors and patterns between waves, and how more infectious virus mutations affect resurgence under strong or weak interventions^4^.

To date, the related research is limited with focus specified on (1) describing the epidemiological characteristics of second waves, e.g., by epidemic renormalization grouping^5^ or stochastic SEIR modeling^6^, on age-sensitive infection and mortality^7–9^, mobility-based regrowth patterns^10^, and risk or trend of resurgence^11–14^; (2) comparing the epidemiological attributes of first and second waves, e.g., younger patients with lower hospitalization duration and fatality rate in second waves^15–17^; (3) studying the impact of NPIs, vaccination and/or virus mutation on transmission and infection^18–22^; and (4) forecasting resurgence resulted from specific external factors such as easing interventions^23–25^, intervention fatigue^26^, physical distancing and vaccination^27^, quarantine, testing and contact tracing^28^, non-pharmaceutical public health intervention^29,30^, social distancing and loss of immunity^31^, virus mutation^32,33^, air pollution^34^, vaccination^35–39^, and disease dynamics and social processes^40^.

Here, we systematically quantify the influence of sequential control and relaxation intervention strategies (including public activities) at different severity and timing and virus mutations at various transmissibility levels on people at different continents and with or without vaccination in epidemic processes and on COVID-19 wave differences and resurgence over 2020 and 2021^2^. By integrating epidemiological knowledge, data-driven discovery, dynamic interaction and event analysis, evidence and findings are extracted from this domain-, data- and event-driven dynamic interaction and influence modeling of COVID-19 first-to-later wave cases, interventions, social activities, and virus mutations. We answer several fundamental questions about COVID-19 resurgence: (1) How do epidemiological attributes change over the continuous resurgences in countries holding different ethnic and cultural backgrounds, intervention strategies, wave patterns, virus mutation stages over 2020 and 2021^3^, and vaccination conditions? (2) How do different severity, number and timing of interventions and public activities individually and cumulatively affect the behaviors and trends of different waves and resurgences? (3) How would different control and relaxation intervention strategies influence the next 30-day trends following a resurgence under substantially vaccinated or unvaccinated conditions? (4) How would more infectious virus mutations like delta or even much worse than delta influence the resurgences and the next 30-day trends under control and relaxation interventions and with substantially vaccinated or unvaccinated conditions?

Accordingly, we develop a dynamic event-driven epidemiological model iSPEIQRD. Here *events* broadly refer to any control or relaxation interventions implemented by governments and activities of individuals or communities that may influence the dynamics of COVID-19 population states and coverage and the epidemic processes, transmissions and trends. iSPEIQRD models event interactions and their impact on epidemiological states, state transition and attributes of population, and epidemic processes. We infer both epidemiological and event-sensitive attributes of the epidemic, interactions, and impact. The epidemiological attributes are the infection rate, the incubation rate, the recovery rate, and the mortality rate, where recovery and mortality rates are time-dependent. The event-sensitive variables are the quarantine rate, the inferred protection rate, and the deconfinement rate of interventions and activities, the individual and cumulative impacts of interventions and activities on cases, and the time delay parameter for interventions and activities to take effect, where protection and deconfinement rates and the time delay parameter are intervention/activity timing dependent. We conduct comprehensive quantitative analyses at comprehensive aspects - *systematic* involving complex real-world scenarios including multi-waves and resurgences evolving over 2020 to 2021, virus mutations, strong and weak interventions, public activities, and vaccination conditions; *vertical* between waves in each country over 2020 and 2021; *horizontal* across countries in different continents with different intervention strategies and vaccination conditions; *contrastive* between vaccinated and unvaccinated conditions; *representative* covering typical ethnic and cultural backgrounds and regions in Europe, Middle East, and Asia; *what-if* with intervention and mutation simulations on resurgence; and *future* for the next 10 to 30-day trends to address the above four questions.

## Main Results

We carefully select five countries with representative but different ethnic and cultural backgrounds, COVID-19 intervention policy preferences and strategies, case development patterns, and vaccination conditions for the case study. Our focus is on comparing the 2020 and 2021 pandemic, virus and vaccination conditions since these stages are more critical and representative for future epidemic management. Three European countries Germany, France and Italy were associated with typical but distinct first and second waves in 2020 without vaccination. A Middle Eastern country Israel and an Asian country Japan were with two waves spreading over 2020 and 2021, affected by virus mutants including delta but with different vaccination conditions. Israel as a most representative country in containing COVID-19 is regarded early and highly vaccinated with active mitigation strategies, while Japan was reluctant to both strict containment and vaccination. Below, we summarize the main results and findings addressing the above four questions (more details in the supporting information).

### How do different severity, number and timing of interventions individually affect the trends of the two waves?

We infer and compare the individual impacts of sequentially implemented intervention, vaccination and response events on cases and patterns of different waves, as shown in Figure 1. By aligning the scale of daily new cases and individual event impact, the impact rate of each event (the time delay factor has already been considered) explains the intervention effectiveness in controlling or relaxing infection. The two waves in each country demonstrate different wave patterns and dynamics as a result of their different severity, number and timing of interventions (including vaccination if any) and public activities (as well as other environmental factors). The events enforced in the first waves had higher containment impact than those implemented in the second waves. Control and relaxation events generate opposite effects, either suppressing or speeding up wave curves. The inappropriate relaxation such as early relaxation and late intervention may have contributed to the wave differences and COVID-19 resurgence.

**Figure 1.**
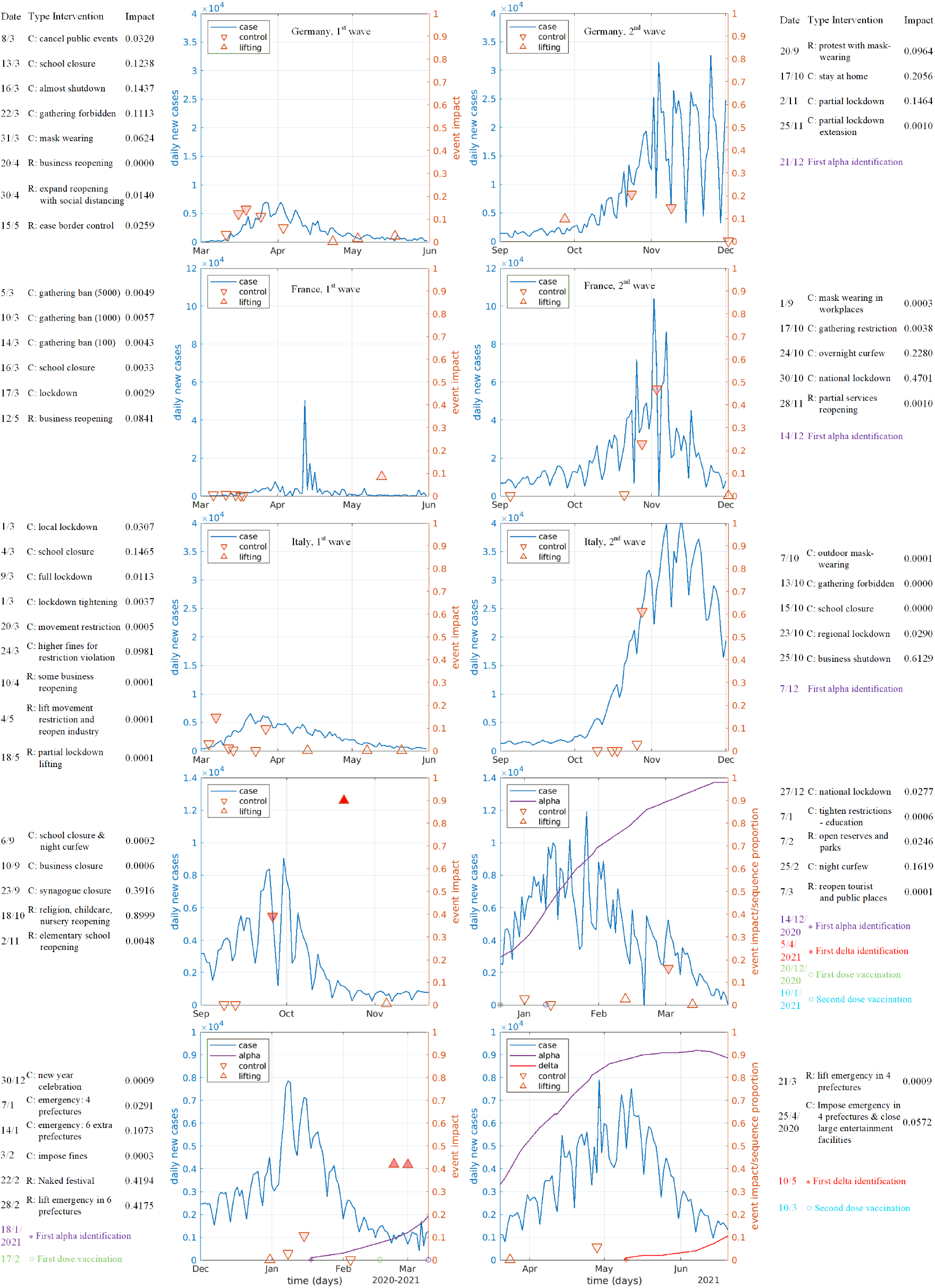
The individual impact of control and relaxation interventions on daily new cases during the two waves in Germany, France, Italy, Israel, and Japan. The blue axis in the diagrams represents the daily new cases, and the red axis represents the event impact rate. The blue curve represents the change of daily new case numbers, the red regular triangles represent relaxation events, and the red inverted triangles represent control events. The deeper the color of the red triangles, the greater the impact of the events on the daily cases. The date, type, description, and impact of each intervention are listed next to the first or second wave’s diagram with the inferred impact rate of each event. The side information shows each event and its impact, the alpha or delta appearance, and the first and second dose vaccination if available.

In **Germany**, the first wave was quickly contained by five powerful control events (including school closure with impact rate 0.1238, almost lockdown at 0.1437 and gathering forbidden at 0.1113) implemented in the initial stage when the cases were low. In the second wave, there is a longer time delay for the events to take effect. A large protest without wearing masks and maintaining social distancing (impact rate 0.0964) happened at the beginning, causing deconfinement and promoting the resurgence. When the cases were already high, two strong interventions (stay at home with impact rate 0.2056 and partial lockdown at 0.1464) did not effectively contain the infection quickly, leading to or associated with a longer period, higher peak value and more cases. The interventions almost shutdown (with impact rate 0.1437) and stay at home (0.2056) had the most significant roles in the first and second waves, respectively.

In **France**, five strong control interventions (including school closure and lockdown) implemented in the initial stage of the first wave when the cases were very low generated an immediate strong impact on containing cases. In the second wave, two initial interventions (mask wearing 0.0003 and gathering restriction 0.0038) implemented when the cases were already high (much higher than the first wave) were too mild to contain the infections. The cases continuously increased and the outbreak lasted much longer with a much higher peak cases. This direction did not change until two strict interventions curfews (0.2280) and lockdown (0.4701) substantially compressed the growth to decline. A longer period with more infections and higher peak cases was seen in the second wave compared to the first.

In **Italy**, the early and strict interventions (local lockdown and school closure) adopted before the daily new cases reached 1,000 effectively contained the first outbreak. In the second wave, effective interventions (business shutdown with impact rate 0.6129) were not taken until the daily new cases exceeded 20,000. Four lagged interventions (including school closure with impact rate 0 and regional lockdown at 0.029) after the cases grew to a substantial level did not take effect. The cases still grew exponentially, which was only compressed after implementing business shutdown at a very high case level.

In **Israel**, the resurgence between Sept. and Nov. 2020 was contained by increasingly enhanced mitigation from locking down the ‘red’ communities to national lockdown and synagogue closure. However, though highly vaccinated, the follow-up full reopening incurred the sharp and dense increase of cases and another resurgence between Dec. 2020 and Apr. 2021. This more volatile wave suffered from less firm interventions, which were mixed with relaxations, and probably the more infectious alpha strains, before it was effectively contained.

**Japan** saw an immediate resurgence with cases doubled after allowing its new year celebration in Jan. 2021. Two follow-up emergency states plus fines on restriction violations pushed down the cases to end the first wave from Dec. 2020 to Mar. 2021. However, the restriction release for celebrating the Japanese Naked Festival and canceling emergency states in Mar. resulted in another resurgence from Apr. to Jul., which was slowly contained by enforcing another emergency state. Though vaccinated, the second wave saw much higher and denser cases than its early wave, likely owing to much less and weaker interventions and more infectious alpha and delta virus strains.

### How do different severity, number and timing of interventions cumulatively affect the trends of the two waves?

We infer and align the cumulative impact of all events and their protection and deconfinement effects on the daily new cases of the first and second waves in Figure 2, where the double-dose vaccination rates for Israel and Japan are also shown as a reference. The overall impact reflects the control vs. relaxation effect of all sequentially implemented events on the daily case movement, wave patterns and differences between two waves. The protection and deconfinement rates measure the incremental effect of all step-wise events on converting people to protected or susceptible states.

**Figure 2.**
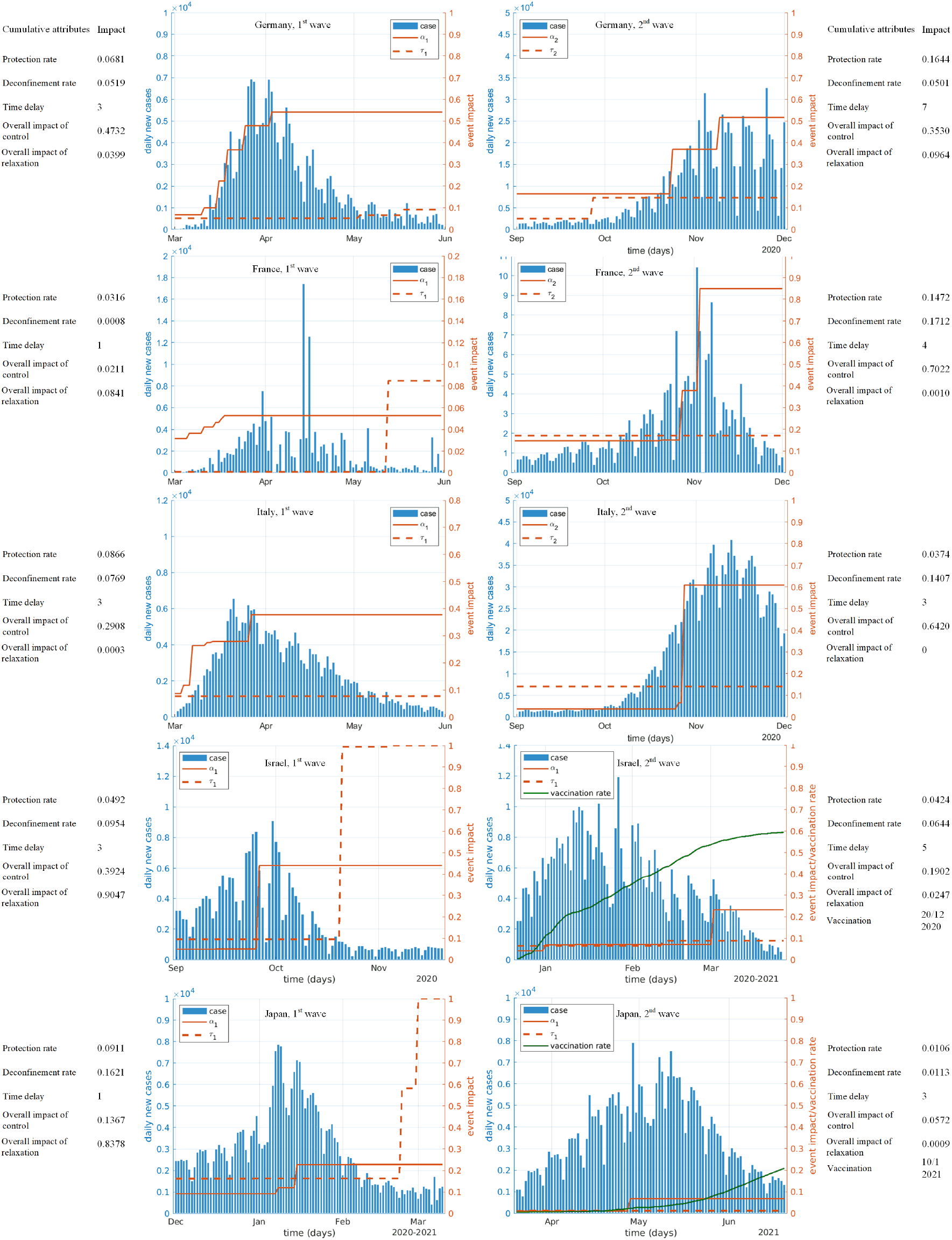
The overall impact and protection-deconfinement effects of sequential interventions on daily new cases during the two waves in five countries. The blue axis represents the daily new cases, and the red axis represents the cumulative impact of sequentially enforced events. The blue bars represent the daily new cases, the red solid line represents the cumulative impact of control events (i.e., the change of protection rate), and the red dash line represents relaxation events (i.e., the change of deconfinement rate). The sequentially implemented control and relaxation events lead to the increase of protection and deconfinement, respectively. The side information shows the inferred rates of protection vs. deconfinement and control vs. relaxation. The green curves refer to the full vaccination rates in Israel and Japan as a reference.

In **Germany**, two waves have similar and low-level initial deconfinement rates (0.0519 vs. 0.0501), reflecting their similar average populations returning from the protected to the susceptible (e.g., losing immunity and returning to workplace, etc.) compartments at the beginning of the two waves when there were no other relaxation events. Contradictory response strategies were implemented during the two waves, producing different effects on case movements as shown by the protection rates (0.0681 vs. 0.1644). Early and strict interventions implemented in the early stage of the first outbreak generated an immediate and strong protection effect and flattened the curve. Relaxation measures were then made after the epidemic was substantially contained, and the deconfinement rate remained low in the whole first wave. In the second wave, the protection effect of the late interventions was highly discounted. The protest led to the increase of the corresponding deconfinement rate and caused the exponential increase of daily new cases until the control events stay at home, partial lockdown and partial lockdown extension took effect, where stay at home and partial lockdown lifted the protection to a significant level and suppressed the curve rising. The first wave shows a higher overall control effect (impact rate at 0.4732) than the second wave (0.3530), but a lower overall relaxation impact (0.0399 vs. 0.0964).

In **France**, the step-wise early and strong interventions in the initial stage of the first wave when the cases were very low resulted in a very low deconfinement rate (0.0008) but a much higher protection rate (0.0316), which effectively contained the first wave. In the second wave, the initial protection rate, the deconfinement rate, and the time delay before events taking effect were larger than that in the first wave. The increased initial deconfinement rate of the second wave may be due to the early relaxation of social restrictions. Then the French government reopened businesses from 12 May. This significant early relaxation may have led to the increase of the initial deconfinement rate in the second wave (as shown by the high cases at the end of the first wave). The late and fewer strong interventions when the cases were approaching their peak values contributed to a much lower protection rate but a much higher deconfinement rate than the first wave, where deconfinement showed stronger impact than protection. Two soft interventions mask-wearing and gathering restrictions resulted in the higher deconfinement rate than the first wave. The overall impact of control interventions in the second wave (at 0.7022) was much larger than the first wave (at 0.0211) since they took effect only until the curve reached its peak. This explains why a broader and larger population were infected in the longer second wave than the first wave.

In **Italy**, the protection rate in the first wave growed quickly and highly exceeded the deconfinement rate. In the initial stage of the second wave, the deconfinement rate was higher than the protection rate, indicating much relaxed control on the epidemic and leading to an exponential growth of daily new cases. The initial protection rate decreased from 0.0866 in the first wave to 0.0374 in the second wave, while the initial deconfinement rate increased by nearly twice. This is different from Germany and France, explaining that the first wave was better managed than the second wave. These parameters conform to their corresponding interventions. Six control events and three relaxation events quickly controlled the first wave, compared to the five events implemented in the second wave only for containment after a big surge of infections, resulting in a softer effect. Accordingly, only strict interventions such as full lockdown could contain the epidemic, while it would take a longer period and cause more infections since it was implemented too late.

In **Israel**, the protection rate substantially exceeds the deconfinement rate at the peak of the first wave, reflecting the conversion from loose to harsh control, which resulted in the containment of the first wave. However, the deconfinement rate was close to 1 at the end of the wave, triggering significant resurgence started in Dec. 2020. Over most of the period of the second wave, the protection and deconfinement share similar strength, indicating weak and relaxed interventions, resulted in higher and denser infections, although vaccination was highly adopted.

In **Japan**, the balance between weak protection and strong deconfinement explains the long and high infection period between Dec. 2020 and Mar. 2021. A similar high deconfinement rate appeared at the end of the first wave, which triggered the resurgence between Mar. and Jul. 2021. While partially vaccinated, the higher and more lasting infections in this wave were attributed to its very limited and more relaxed mitigation, as shown by the balanced protection and deconfinement rates.

### How would different control and relaxation intervention strategies influence the next 30-day trends following the second waves?

We estimate the cases in the next 30 days following the last day of the second wave under six what-if scenarios with either strict control measures or relaxing social recovery, as shown in Figure 3. We vary the protection rate (*α′*) and the deconfinement rate (*τ′*) to simulate six intervention strategies of containing or tolerating COVID-19.

**Figure 3.**
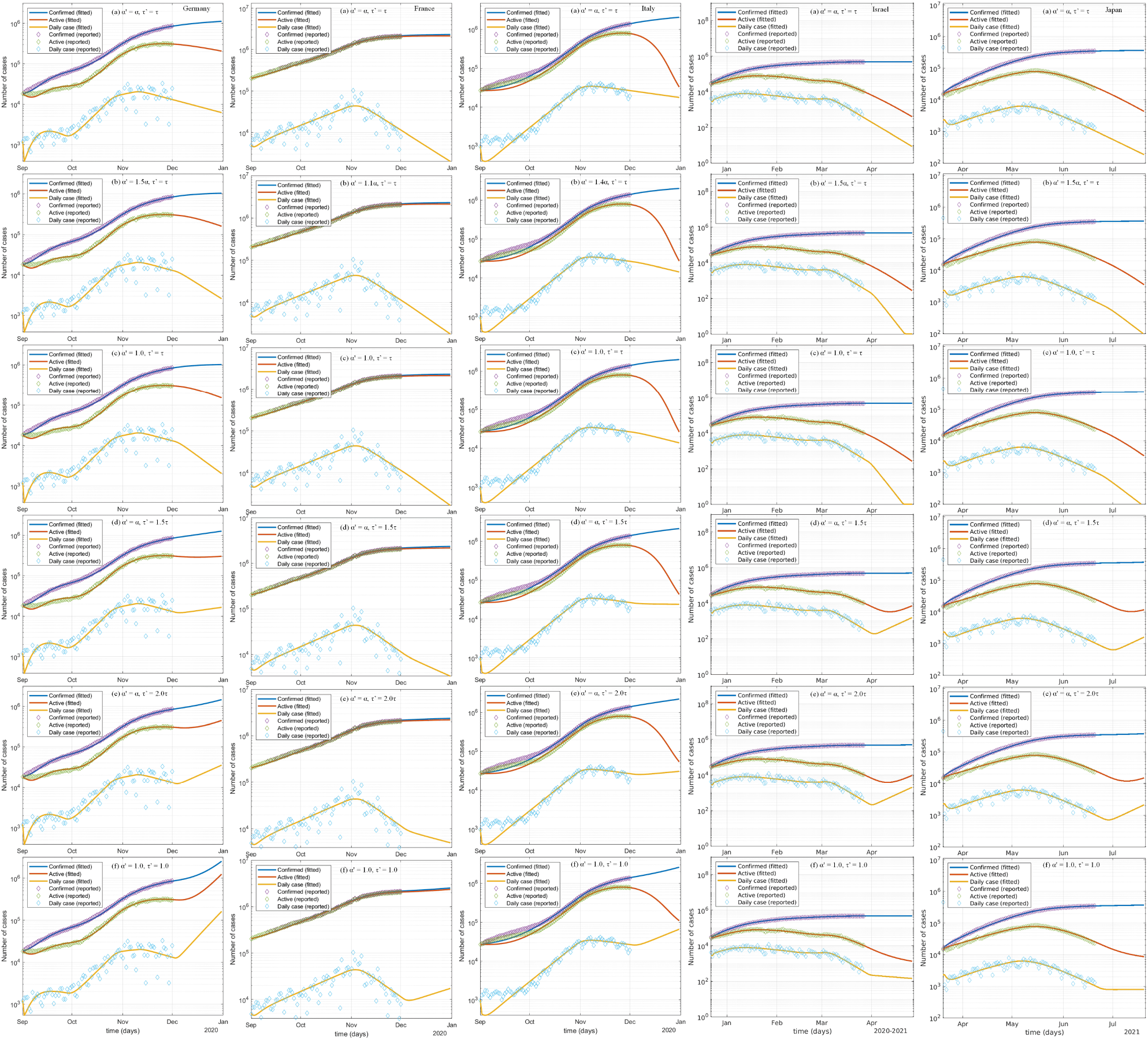
The impact of changing the protection-deconfinement tradeoff by relaxing or enforcing interventions on the next 30-day infections following the second COVID-19 waves in Germany, France, Italy, Israel, and Japan.

First, if the current control interventions remain unchanged, i.e., scenario (a) in Figure 3, the daily new cases would continue to decline in the following 30 days. The wave would be contained by the existing weak interventions allowing certain socioeconomic recovery activities. Second, if the protection rate increases (e.g., by 50% as shown in Germany, Israel and Japan, 10% in France, and 40% in Italy in scenarios (b)) or reaches its maximum 1.0 (i.e., the scenario (c)), more susceptible people would be protected and the epidemic would be contained more quickly. Third, when the protection rate remains unchanged and we only increase the deconfinement rate by 50% (i.e., the scenario (d)) or 100% (i.e., the scenario (e)), these relaxation interventions would result in different trends of the daily new cases in these countries. In Germany, Israel and Japan, the curves continue to decrease in a short time period and then turn to an upward trend.

A larger deconfinement rate implies a faster growth of daily new cases. Undoubtedly, there would be another peak of the daily new cases in scenarios (d) and (e) in Germany, Israel, and Japan. In France, the daily new cases would continue to decrease but the decrease would slow down. This is because, compared with the protection rate, the deconfinement rate at the end of the second wave is quite low (*τ* = 0.1722), even when it doubles (i.e., *τ′* = 2.0*τ* = 0.3444). The epidemic would still be contained by the control interventions and a larger protection rate. A small growth of the deconfinement rate would not affect the trend of the daily new case curve. However, in Italy, the curve would decrease first and then become stable when the deconfinement rate increases by 50% in scenario (d). When it increases to *τ′* = 2.0*τ*, the daily new cases would start to increase after a short-term drop, as shown in scenario (e).

Lastly, we evaluate a balanced strategy by increasing both the protection rate and the deconfinement rate, as shown in scenario (f), to control the epidemic but resume social activities at the same time, such as by reviving societies under strict interventions like social distancing and mask wearing. Accordingly, when we increase the two rates to their maximum values for Germany, France and Italy, their daily new case curves would go upwards quickly after a short-term decrease in the first few days. In contrast, the overall decline in Israel and Japan would substantially slow down with chances to increase. Although vaccinations were adopted, both Israel and Japan show no distinguishable trends from Germany, France and Italy.

Figure 3 also shows the effect of adjusting the six scenarios on the confirmed cases and the daily active cases of the next 30 days. The confirmed cases increase in all scenarios. The change of daily active cases results from the daily new cases, the recovery rate, and the mortality rate, showing different trends in these countries.

### How would more infectious virus mutants influence the second waves under strong or weak interventions?

First, when the infection rate remains unchanged (i.e., no more infectious mutants), the daily cases over the second wave would decrease dramatically if the interventions in the first waves were implemented, as shown in Figure 4(a) in Germany and Italy. This means, though the relaxation in the second wave led to unexpected spread, much stronger interventions would substantially control the resurgences. In France, the daily new case curve did not show a significant deviation from the reported until the turning point at the beginning of November. The large increase of cases leads to a great gap and difference between the two waves in France and the simulation shows that, even if the same interventions as the first wave were implemented in the second wave, the control intensity would still be insufficient to quickly and effectively contain the severe resurgence, probably requiring stronger and earlier interventions. In Israel, the infections would increase and then drop significantly in middle January, while the trend would change in middle Feb. significantly again. This shows the carried interventions implemented in the early stage (i.e., three increasing lockdowns up to end of September) of its early wave would be strong enough to contain the virus. However, transferring the strategies of easing lockdown and reopening schools in Oct. and Nov. to the second wave would significantly lift the cases since middle Feb. In Japan, the interventions (including the emergency state in Jan.) implemented in the first wave are more intensive than that in the second resurgence, transferring them to the second wave would overall contain the infections well, including changing the wave direction in middle Mar. However, transferring two strong relaxing activities (Naked festival and lifting emergency state) at the end of the first wave would accordingly change the infection trend at the end of the second wave.

**Figure 4.**
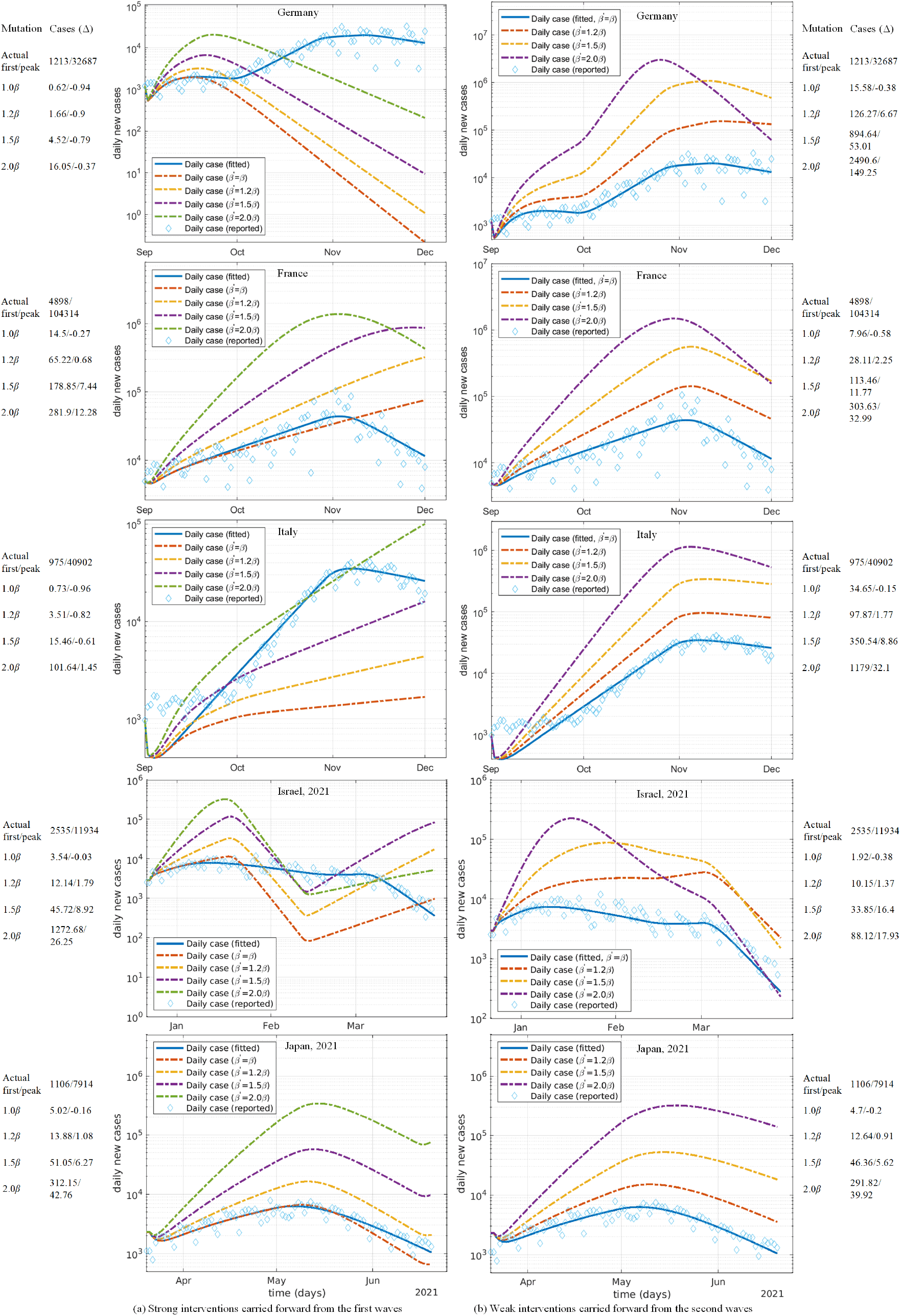
The impact of more infectious virus mutants on the second wave under (a) strong interventions (i.e., the same control interventions as in the first wave), and (b) weak interventions (the same relaxed interventions as in the second wave). The side information shows the times of change of estimated peak daily cases over the actual cases of the first and peak days.

Second, we assume more infectious virus mutants appeared in the second waves by increasing the transmission rates by 20%, 50% and 100% (i.e., *β′* = 1.2*β, β′* = 1.5*β* and *β′* = 2.0*β*)^4^. We then simulate their effects on the daily cases by transferring the same interventions from the first to the second waves. Figure 4(a) shows the daily cases under these three mutation scenarios after directly applying the stronger interventions from the first waves. Similar trends would follow the above unmutated transfer, higher infections would be observed in all countries, and more transmissible mutants like delta and lambda would substantially increase the daily cases. For example (Table 18), in Germany, after 2-3 weeks, the daily new cases with the three mutations would reach their peaks at 3,226, 6,691 and 20,677, i.e., increasing 1.7, 4.5 and 16 times compared with the actual observed daily new cases on the first day. The daily cases with the three mutants would decrease to 1, 9, and 208 at the end of the second wave. The more contagious mutations would be eventually controlled after making stronger interventions. In France, a second wave with a more infectious virus would see many more cases. A 50% transmissibility increase would lift the peak cases by 7 times to 880,925 cases, while a 100% infection rate increase would increase the peak value by 12 times to 1,385,640 cases on the peak day. Much stronger and earlier interventions would be essential compared to Germany and Italy. In Italy, the daily cases would reach 4,397 for 1.2*β* (decreasing by 82%) or 16,048 for 1.5*β* (decreasing by 61%)) on the peak days but 100,079 for 2.0*β* (increasing 101 times from that on the first day and 1.45 times than the actual peak cases). In Israel, increasing the mutation by 20% would result in 12 times and about 2 times of increase of cases over the first day and peak day of the second wave, respectively. In contrast, an 50% increase of mutation would result in 45 times and 9 times of case growth; and 100% mutation increase would bring about 1272 times and 26 times of cases instead. Similarly significant growth of cases would be seen in Japan, for example, an increase of 100% mutation would result in 312 times and 42 times of case increase over their first day and peak day in the second wave.

**Table 1.**
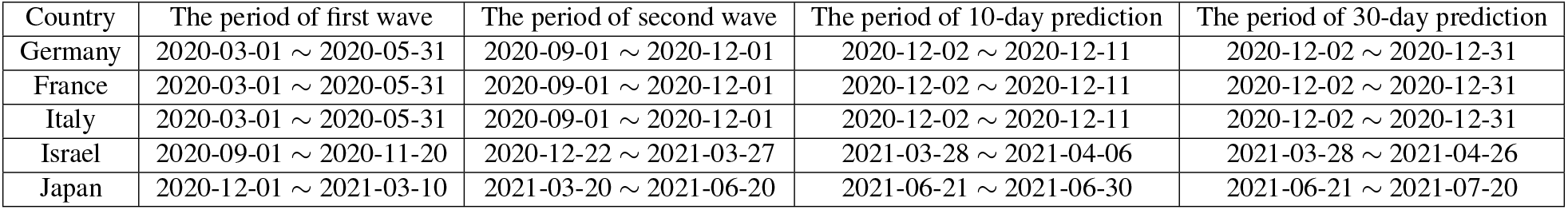
The start and end dates for two waves, 10-day prediction, and 30-day prediction in Germany, France, Italy, Israel and Japan.

| Country | The period of first wave | The period of second wave | The period of 10-day prediction | The period of 30-day prediction |
| --- | --- | --- | --- | --- |
| Germany | 2020-03-01 ~ 2020-05-31 | 2020-09-01 ~ 2020-12-01 | 2020-12-02 ~ 2020-12-11 | 2020-12-02 ~ 2020-12-31 |
| France | 2020-03-01 ~ 2020-05-31 | 2020-09-01 ~ 2020-12-01 | 2020-12-02 ~ 2020-12-11 | 2020-12-02 ~ 2020-12-31 |
| Italy | 2020-03-01 ~ 2020-05-31 | 2020-09-01 ~ 2020-12-01 | 2020-12-02 ~ 2020-12-11 | 2020-12-02 ~ 2020-12-31 |
| Israel | 2020-09-01 ~ 2020-11-20 | 2020-12-22 ~ 2021-03-27 | 2021-03-28 ~ 2021-04-06 | 2021-03-28 ~ 2021-04-26 |
| Japan | 2020-12-01 ~ 2021-03-10 | 2021-03-20 ~ 2021-06-20 | 2021-06-21 ~ 2021-06-30 | 2021-06-21 ~ 2021-07-20 |

**Table 2.** The dates of identifying the first alpha and delta strains and implementing the first and second dose vaccinations in Germany, France, Italy, Israel and Japan.

| Country | First alpha identification | First delta identification | First dose vaccination | Second dose vaccination |
| --- | --- | --- | --- | --- |
| Germany | 2020-12-21 | 2021-05-10 | 2020-12-27 | 2020-12-27 |
| France | 2020-12-14 | 2021-05-03 | 2020-12-27 | 2021-01-16 |
| Italy | 2020-12-07 | 2021-05-15 | 2020-12-27 | 2021-01-04 |
| Israel | 2020-12-14 | 2021-04-05 | 2020-12-20 | 2021-01-10 |
| Japan | 2021-01-18 | 2021-05-10 | 2021-02-17 | 2021-03-10 |

**Table 3.** The major interventions and activities reported during the two COVID-19 waves in Germany.

| Date | Event | Type |
| --- | --- | --- |
| 08 Mar. | cancel public events of more than 1,000 attendees | C |
| 13 Mar. | close schools and nurseries | C |
| 16 Mar. | implement almost shutdown | C |
| 22 Mar. | forbid gatherings | C |
| 31 Mar. | encourage mask wearing | C |
| 20 Apr. | start to reopen shops | R |
| 30 Apr. | expand reopening under strict social distancing conditions | R |
| 15 May | ease border controls | R |
| 03 Jun. | allow travel to all 26 EU countries | R |
| 13 Jun. | lift most contact restrictions | R |
| 01 Aug. | have a 20,000 people protest ignoring mask-wearing and physical distancing requirements | R |
| 20 Sep. | have a protest with mask-wearing requirements | R |
| 17 Oct. | start staying at home and contact tracing | C |
| 02 Nov. | start partial lockdown | C |
| 25 Nov. | extend partial lockdown, etc. | C |

**Table 4.** The major interventions and activities reported during the two COVID-19 waves in France.

| Date | Event | Type |
| --- | --- | --- |
| 05 Mar. | ban on gatherings of more than 5,000 people | C |
| 10 Mar. | ban on gatherings of more than 1,000 people | C |
| 14 Mar. | ban on gatherings of more than 100 people | C |
| 16 Mar. | close schools and public establishments | C |
| 17 Mar. | start lockdown | C |
| 12 May | allow business reopening and gatherings per social distancing rules | R |
| 30 May | reopen parks | R |
| 02 Jun. | reopen restaurants and museums | R |
| 15 Jun. | permit demonstrations per social distancing guidelines | R |
| 22 Jun. | reopen cinemas | R |
| 11 Jul. | lift majority of restrictions | R |
| 01 Sep. | require mask wearing in workplaces | C |
| 17 Oct. | start gathering restriction | C |
| 24 Oct. | start overnight curfews | C |
| 30 Oct. | start second national lockdown | C |
| 28 Nov. | reopen non-essential services | R |

**Table 5.**
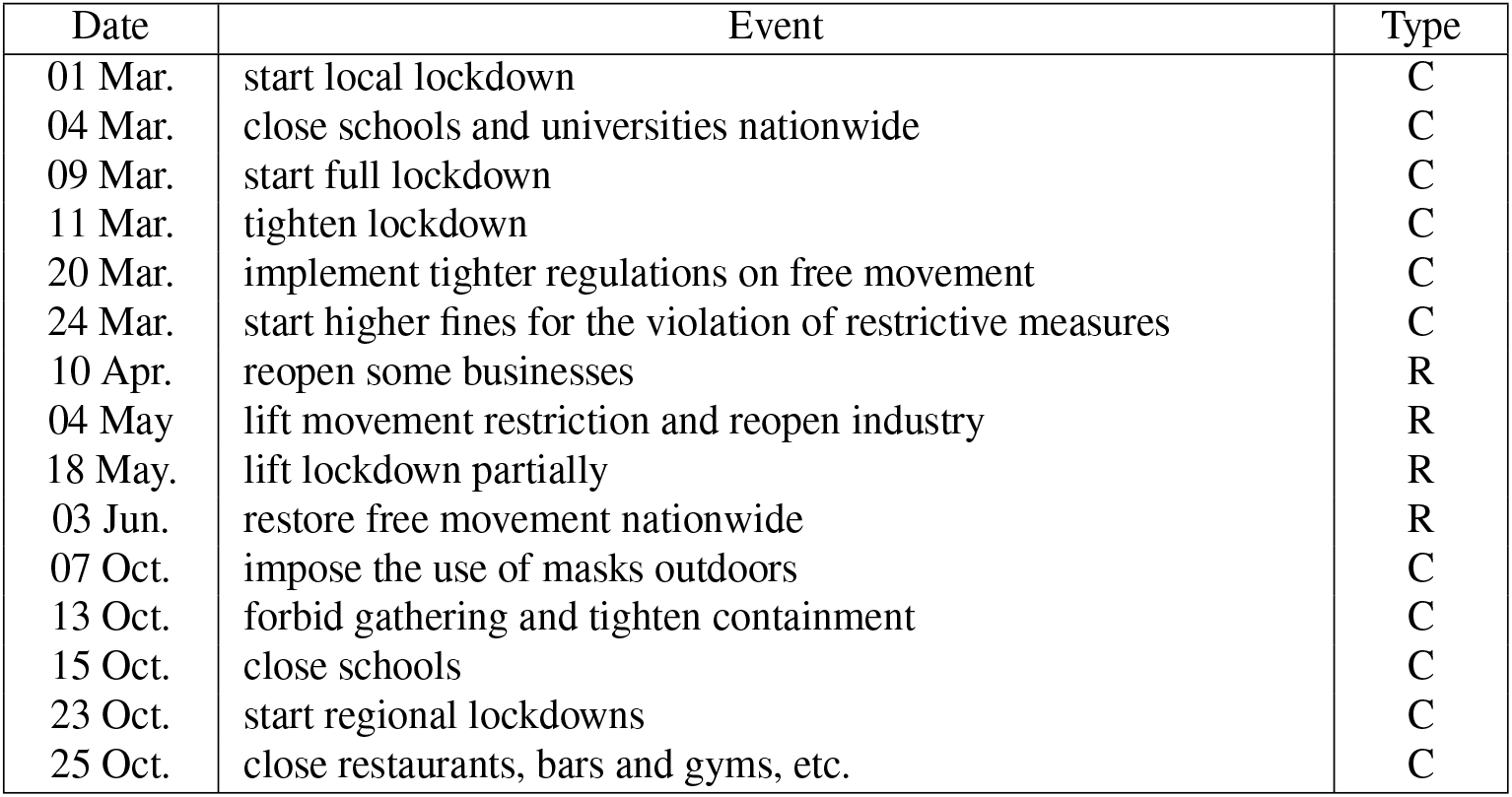
The major interventions and activities reported during the two COVID-19 waves in Italy.

| Date | Event | Type |
| --- | --- | --- |
| 01 Mar. | start local lockdown | C |
| 04 Mar. | close schools and universities nationwide | C |
| 09 Mar. | start full lockdown | C |
| 11 Mar. | tighten lockdown | C |
| 20 Mar. | implement tighter regulations on free movement | C |
| 24 Mar. | start higher fines for the violation of restrictive measures | C |
| 10 Apr. | reopen some businesses | R |
| 04 May | lift movement restriction and reopen industry | R |
| 18 May. | lift lockdown partially | R |
| 03 Jun. | restore free movement nationwide | R |
| 07 Oct. | impose the use of masks outdoors | C |
| 13 Oct. | forbid gathering and tighten containment | C |
| 15 Oct. | close schools | C |
| 23 Oct. | start regional lockdowns | C |
| 25 Oct. | close restaurants, bars and gyms, etc. | C |

**Table 6.** The major interventions and activities reported during the two COVID-19 waves in Israel.

| Date | Event | Type |
| --- | --- | --- |
| 06 Sep. 2020 | school closure and night-time curfew for ‘red’ communities | C |
| 10 Sep. 2020 | national lockdown: mall, store and school closure | C |
| 23 Sep. 2020 | synagogue closure | C |
| 18 Oct. 2020 | ease national lockdown: reopen religious sites, kindergartens and nurseries | R |
| 02 Nov. 2020 | reopen elementary schools (grades 1-4) | R |
| 27 Dec. 2020 | a national lockdown: travel and gathering limit, close all non-essential stores and services | C |
| 07 Jan. 2021 | tighten restrictions: close the entire education system | C |
| 07 Feb. 2021 | ease lockdown: reopen open-air nature reserves and parks | R |
| 25 Feb. 2021 | curfew from 8:30pm to 5:00am | C |
| 07 Mar. 2021 | ease further restrictions: open tourist attractions and public places | R |

**Table 7.** The major interventions and activities reported during the two COVID-19 waves in Japan.

| Date | Event | Type |
| --- | --- | --- |
| 30 Dec. 2020 | Japanese New Year: celebration and events | R |
| 07 Jan. 2021 | emergency state in four prefectures: restrict unnecessary outings after 8pm and close large public places at 8pm | C |
| 14 Jan. 2021 | emergency state in additional six prefectures | C |
| 03 Feb. 2021 | impose fines on restriction violations | C |
| 22 Feb. 2021 | Japanese Naked Festival: gathering and celebration | R |
| 28 Feb. 2021 | lift emergency state in six prefectures | R |
| 21 Mar. 2021 | lift emergency state in four prefectures | R |
| 25 Apr. 2021 | impose emergency state in four prefectures: close large entertainment facilities | C |

**Table 8.**
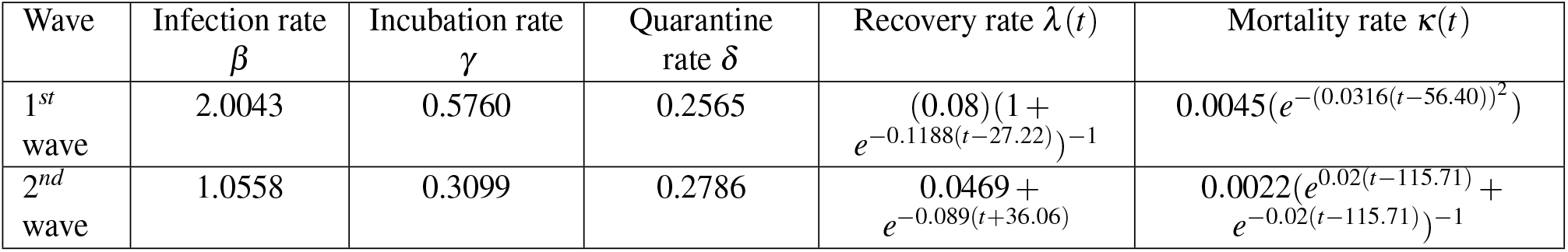
Epidemiological attributes of the 2020 first-to-second waves in Germany.

**Table 9.**
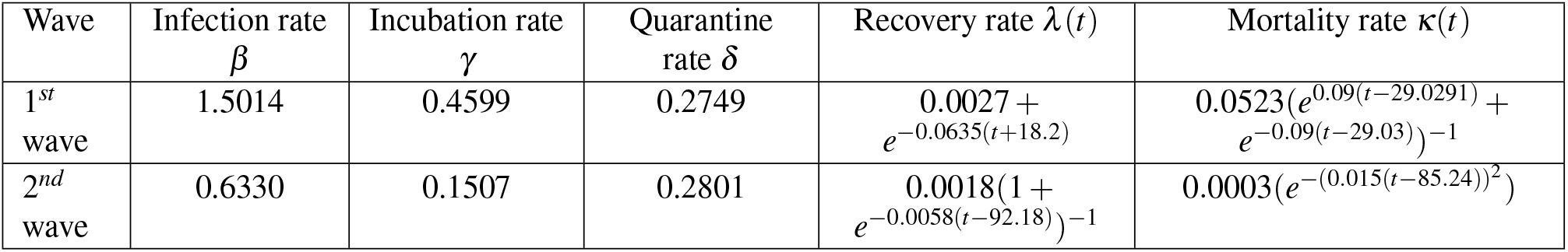
Epidemiological attributes of the 2020 first-to-second waves in France.

**Table 10.**
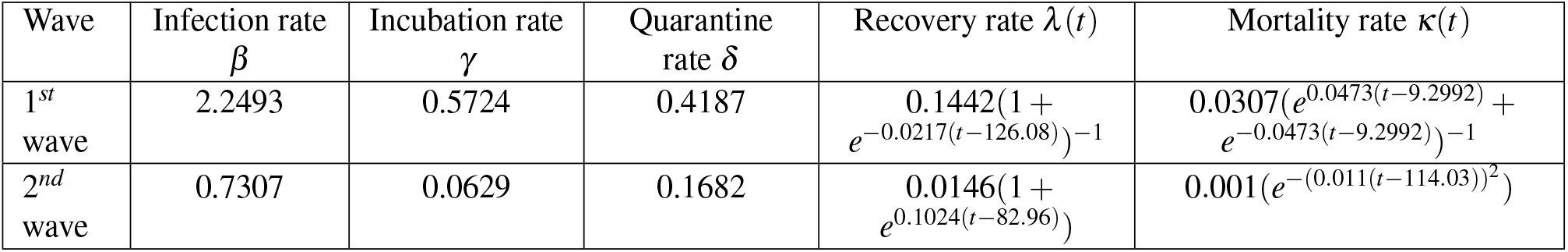
Epidemiological attributes of the 2020 first-to-second waves in Italy.

Lastly, Figure 4(b) shows the simulation results when increasing the infection rate by 20%, 50% and 100% under relatively soft interventions as implemented in the second waves in the five countries, the daily new cases would increase significantly (Table 19). For example, with a 100% transmissibility increase, on the peak day, 3,022,314 Germans (3.8% of their population), 1,492,105 French people (2.2%), 1,150,504 Italians (1.9%), 225,915 (2.6%), and 323,853 (0.3%) would be infected under the same interventions implemented for their second waves, such as stay at home, partial lockdown, mask wearing, gathering restrictions, overnight curfew, and emergency state. Specifically, the peak value would increase 7, 53 and 149 times in Germany, resulted from 20%, 50% and 100% increase of infection. France would see 2, 12 and 33 times of peak case increase. In Italy, the peak value would increase 2, 9 and 32 times due to the stronger transmissibility. In Israel, the peak case number would increase by 1.37, 16, and 18 times. In Japan, there would be 0.9, 5.6 and 40 times of more cases on the peak day of the second wave, as a result of 20%, 50% and 100% transmissibility increase. This analysis (without considering vaccine-induced immunity) also shows no significant impact difference of virus mutations on unvaccinated vs. vaccinated countries.

**Table 11.** Epidemiological attributes of the 2020-2021 first-to-second waves in Israel.

| Wave | Infection rate $\beta$ | Incubation rate $\gamma$ | Quarantine rate $\delta$ | Recovery rate $\lambda(t)$ | Mortality rate $\kappa(t)$ |
| --- | --- | --- | --- | --- | --- |
| 1<sup>st</sup> wave | 1.0465 | 0.2250 | 0.5486 | $0.1199(1 + e^{-0.0559(t-12.57)})^{-1}$ | $0.0016e^{-(0.0282(t-62.22)^2)}$ |
| 2<sup>nd</sup> wave | 1.4207 | 0.3640 | 0.7275 | $0.2893(1 + e^{-0.0078(t-132.16)})^{-1}$ | $0.0006 + e^{(-0.6003(t+74.58))}$ |

**Table 12.** Epidemiological attributes of the 2020-2021 first-to-second waves in Japan.

| Wave | Infection rate $\beta$ | Incubation rate $\gamma$ | Quarantine rate $\delta$ | Recovery rate $\lambda(t)$ | Mortality rate $\kappa(t)$ |
| --- | --- | --- | --- | --- | --- |
| 1<sup>st</sup> wave | 1.4537 | 0.3908 | 0.8188 | $0.2330(1 + e^{-0.0093(t-124.56)})^{-1}$ | $0.0043e^{-(0.0222(t-92.06)^2)}$ |
| 2<sup>nd</sup> wave | 0.7274 | 0.0947 | 0.3581 | $0.3147(1 + e^{-0.0085(t-199.99)})^{-1}$ | $0.0012 + e^{(-0.3050(t+66.76))}$ |

**Table 13.**
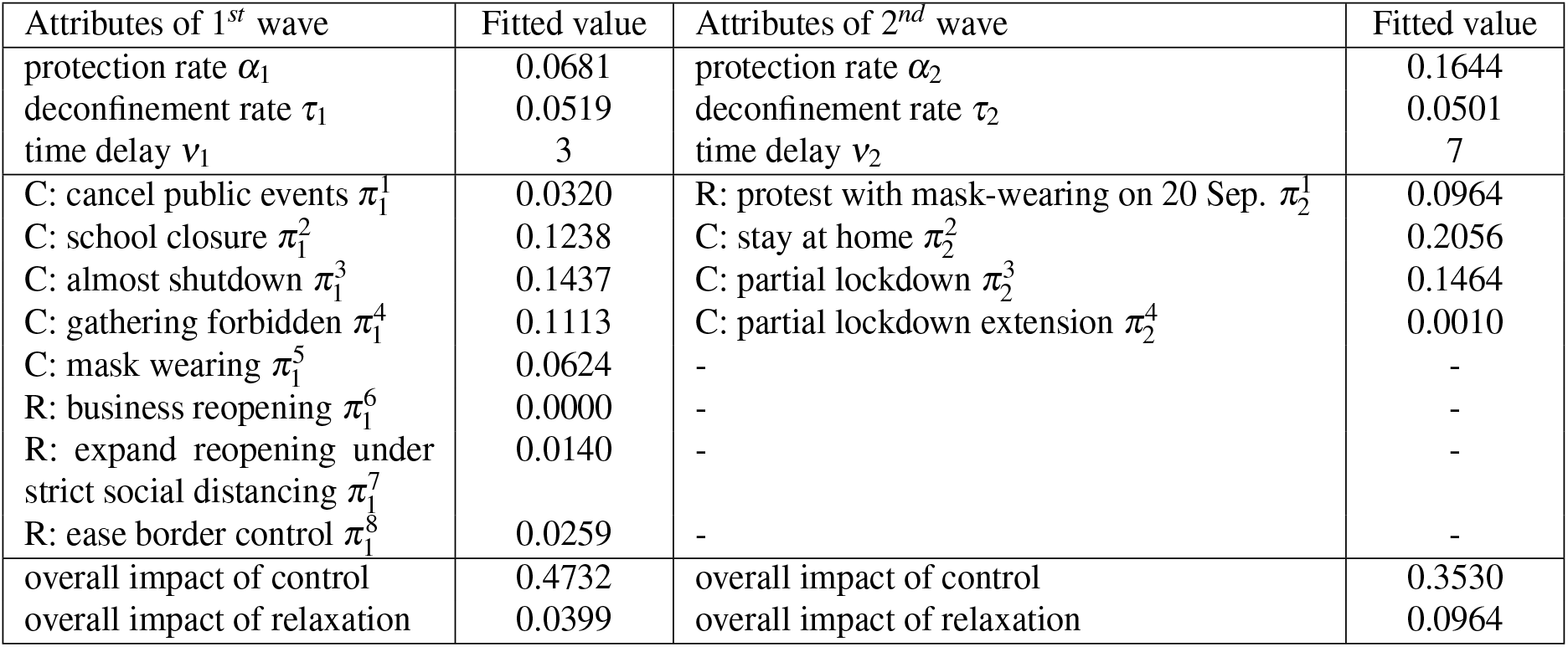
Attributes of event impact of 2020 first-to-second waves in Germany.

**Table 14.** Attributes of event impact of 2020 first-to-second waves in France.

| Attributes of the 1<sup>st</sup> wave | Fitted value | Attributes of the 2<sup>nd</sup> wave | Fitted value |
| --- | --- | --- | --- |
| protection rate $\alpha_1$ | 0.0316 | protection rate $\alpha_2$ | 0.1472 |
| deconfinement rate $\tau_1$ | 0.0008 | deconfinement rate $\tau_2$ | 0.1712 |
| time delay $\nu_1$ | 1 | time delay $\nu_2$ | 4 |
| C: gathering ban (5000) $\pi_1^1$ | 0.0049 | C: mask wearing in workplaces $\pi_2^1$ | 0.0003 |
| C: gathering ban (1000) $\pi_1^2$ | 0.0057 | C: gathering restriction $\pi_2^2$ | 0.0038 |
| C: gathering ban (100) $\pi_1^3$ | 0.0043 | C: overnight curfew $\pi_2^3$ | 0.2280 |
| C: school closure $\pi_1^4$ | 0.0033 | C: national lockdown $\pi_2^4$ | 0.4701 |
| C: lockdown $\pi_1^5$ | 0.0029 | R: partial services reopening $\pi_2^5$ | 0.0010 |
| R: business reopening $\pi_1^6$ | 0.0841 | - | - |
| overall impact of control | 0.0211 | overall impact of control | 0.7022 |
| overall impact of relaxation | 0.0841 | overall impact of relaxation | 0.0010 |

**Table 15.**
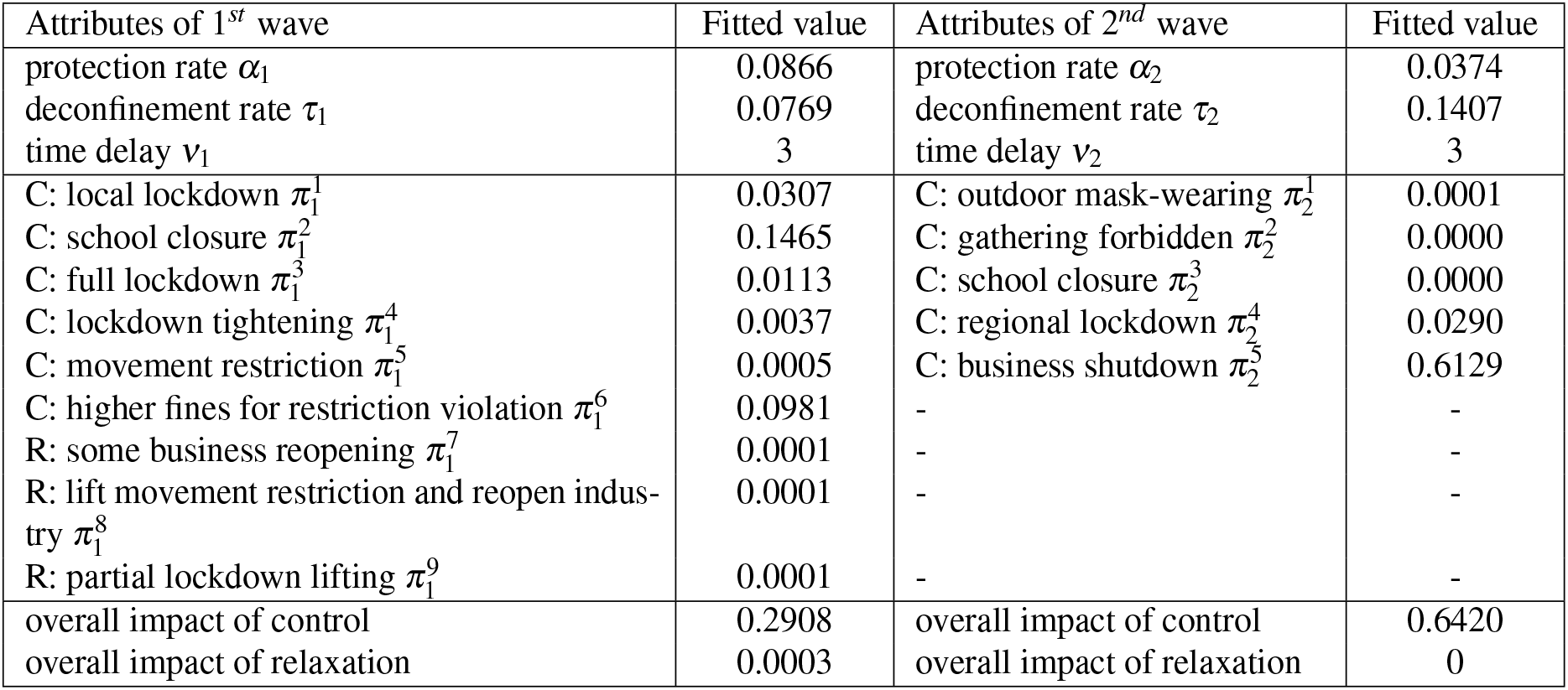
Attributes of event impact of 2020 first-to-second waves in Italy.

**Table 16.** Attributes of event impact of 2020-2021 first-to-second waves in Israel.

| Attributes of 1<sup>st</sup> wave | Fitted value | Attributes of 2<sup>nd</sup> wave | Fitted value |
| --- | --- | --- | --- |
| protection rate $\alpha_1$ | 0.0492 | protection rate $\alpha_2$ | 0.0424 |
| deconfinement rate $\tau_1$ | 0.0954 | deconfinement rate $\tau_2$ | 0.0644 |
| time delay $v_1$ | 3 | time delay $v_2$ | 5 |
| C: school closure and night-time curfew for 'red' communities $\pi_1^1$ | 0.0002 | C: national lockdown: travel and gathering limit, close all non-essential stores and services $\pi_2^1$ | 0.0277 |
| C: national lockdown: mall, store and school closure $\pi_1^2$ | 0.0006 | C: tighten restrictions: close the entire education system $\pi_2^2$ | 0.0006 |
| C: synagogue closure $\pi_1^3$ | 0.3916 | R: ease lockdown: reopen open-air nature reserves and parks $\pi_2^3$ | 0.0246 |
| R: ease national lockdown: reopen religious sites, kindergartens and nurseries $\pi_1^4$ | 0.8999 | C: curfew from 8:30pm to 5:00am $\pi_2^4$ | 0.1619 |
| R: reopen elementary schools (grades 1-4) $\pi_1^5$ | 0.0048 | R: ease further restrictions: open tourist attractions and public places $\pi_2^5$ | 0.0001 |
| overall impact of control | 0.3924 | overall impact of control | 0.1902 |
| overall impact of relaxation | 0.9047 | overall impact of relaxation | 0.0247 |

**Table 17.** Attributes of event impact of 2020-2021 first-to-second waves in Japan.

| Attributes of 1<sup>st</sup> wave | Fitted value | Attributes of 2<sup>nd</sup> wave | Fitted value |
| --- | --- | --- | --- |
| protection rate $\alpha_1$ | 0.0911 | protection rate $\alpha_2$ | 0.0106 |
| deconfinement rate $\tau_1$ | 0.1621 | deconfinement rate $\tau_2$ | 0.0113 |
| time delay $\nu_1$ | 1 | time delay $\nu_2$ | 3 |
| R: Japanese New Year: celebration and events $\pi_1^1$ | 0.0009 | R: lift emergency state in four prefectures $\pi_2^1$ | 0.0009 |
| C: emergency state in four prefectures: restrict unnecessary outings after 8pm and close large public places at 8pm $\pi_1^2$ | 0.0291 | C: impose emergency state in four prefectures: close large entertainment facilities $\pi_2^2$ | 0.0572 |
| C: emergency state in additional six prefectures $\pi_1^3$ | 0.1073 | - | - |
| C: impose fines on restriction violations $\pi_1^4$ | 0.0003 | - | - |
| R: Japanese Naked Festival: gathering and celebration $\pi_1^5$ | 0.4194 | - | - |
| R: lift emergency state in six prefectures $\pi_1^6$ | 0.4175 | - | - |
| overall impact of control | 0.1367 | overall impact of control | 0.0572 |
| overall impact of relaxation | 0.8378 | overall impact of relaxation | 0.0009 |

**Table 18.**
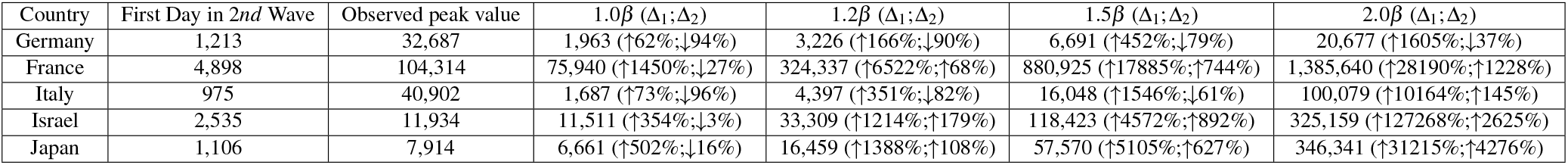
Change of peak daily cases in the second wave caused by more infectious mutants with strong interventions (i.e., the same as in the first waves) in three European countries, corresponding to Figure 4(a). ∆_1_ refers to the change of daily cases on the estimated peak day compared to the first day of the second wave; ∆_2_ refers to the change of daily cases on the estimated peak day compared to the actual peak value of the second wave.

**Table 19.**
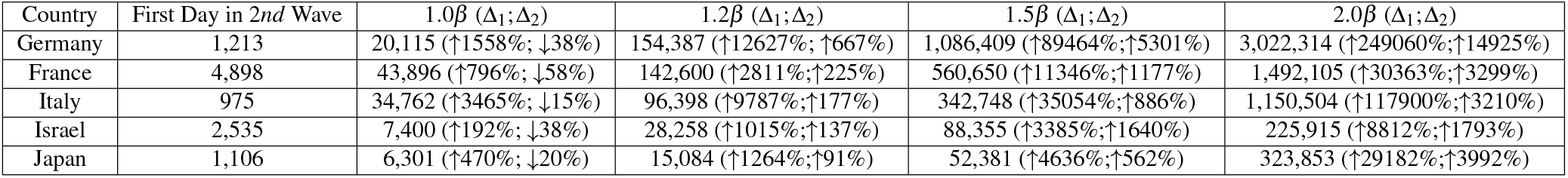
Change of peak daily cases in the second wave caused by more infectious mutants with soft interventions (i.e., the same as in the second waves) in three European countries, corresponding to Figure 4(b). ∆_1_ refers to the change of daily cases on the estimated peak day compared to the first day of the second wave; ∆_2_ refers to the change of daily cases on the estimated peak day compared to the actual peak value of the second wave.

### How would the second waves evolve in the next 30 days with more infectious virus mutants?

We estimate the case movement in the next 30 days following the second wave if virus mutants with different levels of transmissibility would largely spread into unvaccinated and vaccinated communities. Figure 5 shows the estimation results corresponding to three scenarios: mutants with 20%, 50% and 100% infection rate increase, i.e., *β′* = 1.2*β, β′* = 1.5*β*, and *β′* = 2.0*β* in all five countries. The side information shows the actual cases on the last day of the second wave and the mutation-driven proportional variances compared to the last day and the estimated cases at 1.0*β* on the 30^*th*^ day.

**Figure 5.**
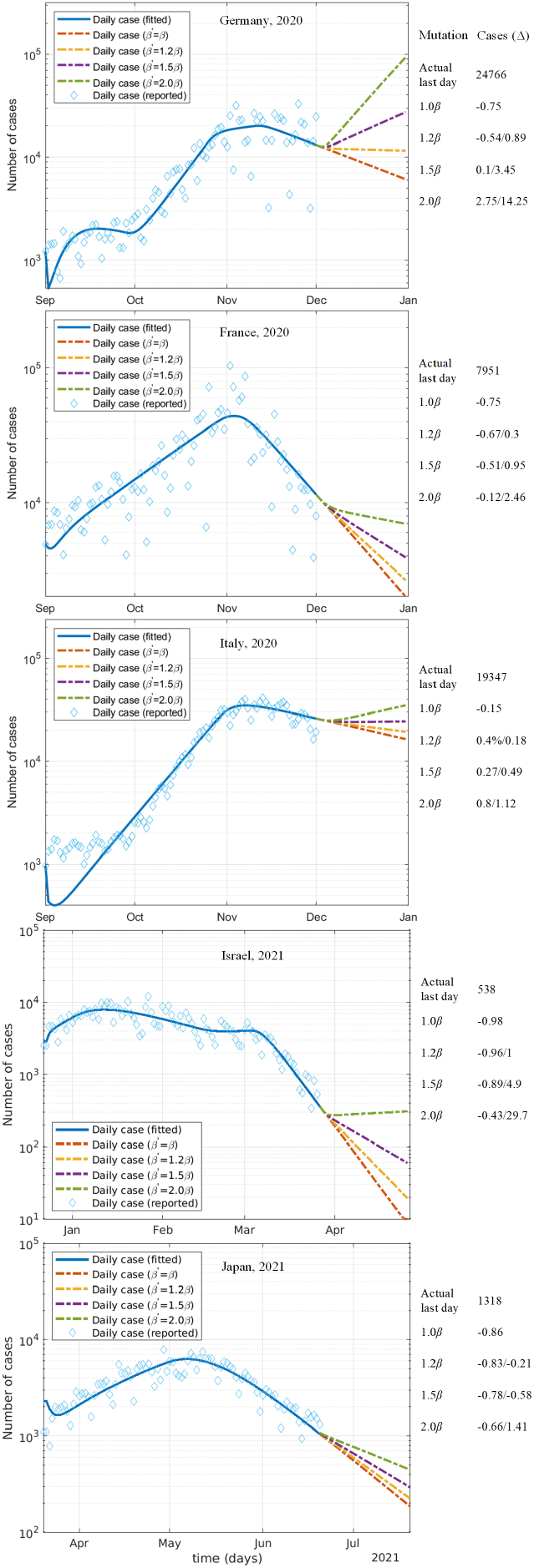
The impact of more infectious virus mutants on daily cases in the next 30 days following the second waves. The side information shows the times of changes of estimated cases over the cases of the actual last day.

As a result of introducing more infectious viruses, the daily cases would significantly increase in Germany, France, and Italy if no vaccination was available, making control even harder and longer. The simulation (Table 20) shows that the mutants with 20%, 50% and 100% extra transmissibility would lead to 11,516 (54% decrease compared to the last day of the second wave), 27,128 (10% increase) and 92,825 (3 times increase) in daily new cases on the 30^*th*^ day in Germany, building on the same other attributes of the original virus. In France, on the 30^*th*^ day, there would be 2,661 daily new cases (30% increase over the estimate for the same day without transmissibility increase i.e. at the 1.0*β* infection rate), 3,915 (95% increase) and 6,962 (2.5 times increase) compared to an estimate of 19,415 (0.4% increase over the last day of the second wave), 24,487 (27% increase) and 34,858 (80% increase) in Italy, as a result of increasing the transmission ability through more infectious mutants. The impact of mutations on the substantially vaccinated Israeli community is not proportionally distinguishable from unvaccinated Germany, France and Italy. While Israel held the world-best vaccination conditions at that time, without considering vaccine-induced immunity, the virus with 100% transmissibility increase would grow the cases by 30 times in comparison with no mutation and significantly reduce the decline trend, much higher that in Germany (14.25 times), France (2.46 times), and Italy (1.12 times). In Japan, where vaccination has been enforced as well while its rate is very low, the 100% more infectious virus would increase the cases by 1.4 times compared to the scenario without mutation and would reduce the decline trend as well.

**Table 20.**
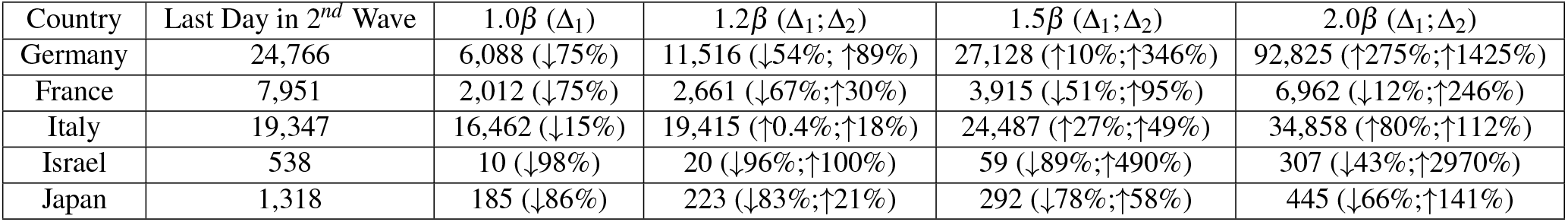
Change of the estimated daily cases on the 30^*th*^ day after the second wave caused by more infectious mutants in three European countries, corresponding to Figure 5. ∆_1_ refers to the change of estimated daily cases on the 30^*th*^ day compared to the last day of the second wave; ∆_2_ refers to the change of estimated daily cases compared to the estimated cases at 1.0*β* on the 30^*th*^ day.

As shown in Germany, Italy and Israel, if a mutant doubles its transmission rate, then it would change the trend of daily new cases from a downtrend to an upward trend. Although the simulation results in France and Japan look relatively optimistic, similar to other countries, their second waves would last much longer with significantly more cases. It would be much harder to control the spread with their mild interventions. This shows the potential overwhelming outcomes of highly infectious coronavirus mutations in the community where no effective control measures are adopted and vaccination is insufficient to contain the virus.

## Discussion

This quantitative analyses provide evidence and indication for the following findings and insights in containing COVID-19.

First, regarding wave differences: (1) Better contained first waves with different wave patterns: all countries applied many control interventions and quickly contained the infection. Germany and Italy had a similar long-tailed movement, as a result of their late implemented interventions after the outbreak became severe and the early relaxation of some interventions. France implemented and retained several less powerful interventions at the very beginning of the first wave, resulting in a balanced single-peak distribution of cases. In Israel, the long tail of its early resurgence was replaced by higher and denser infections in the second resurgence. Similar higher and denser infections were seen in the second resurgence than its first in Japan. (2) Longer and higher second waves: The second waves show more differences than the first ones. Germany and Italy share a more similar distribution with a longer period but a lower number of infections than France, resulting from different interventions. In comparison to Germany who applied mild interventions (partial lockdown), France and Italy enforced interventions with one to multiple orders of magnitude stronger impact (e.g. national lockdown) to effectively control the outbreak. While Israel and Japan adopted vaccination, their second waves were higher with more infections. In all countries, the second waves show higher peak cases than their first waves. (3) Different severity of interventions on wave differences: The lagged enforcement of strongly effective interventions had a prominent impact on the second waves. However, the impact of similar events differed under the distinct situations of two waves. As partial to full lockdown is often chosen as a preferred (or even first choice) effective intervention in both waves, it is critical for policymakers to choose the mostly effective interventions at the right time and duration for the expected consequences. (4) Different timing of interventions on wave differences: While relaxing restrictions addresses the negative public sentiment and socioeconomic recovery, wrong timing (too early or too fast) particularly relaxing controls too soon or enforcing control too late rapidly increases cases and extends the resurgence.

Second, regarding the severity, timing and protection-deconfinement tradeoff of control and relaxation interventions: (1) Intervention severity: Strong interventions such as stay at home, school and business closure, partial to full lockdown, overnight curfew and high fines for restriction violation contribute to a high proportional impact, while weak interventions like mask wearing, social distancing and gathering forbidden result in low impact. (2) Intervention timing: Early strong interventions likely result in instant containment with a shorter, lower and narrower wave of infections. Late strong interventions may extend the infection to a longer period with more cases and high peak cases. (3) Individual impact: Each intervention contributes different proportions to influence the overall case movement (Tables 13-15). A strong intervention may not work to the same proportional impact level when it is applied at different timing in relation to the curve trend. A relaxation event may not incur resurgence if it is implemented in the tail of the decline part of a curve. (4) Cumulative impact: All sequential interventions interact with each other and jointly contribute to an epidemic containment (Tables 13-15). (5) Protection-deconfinement tradeoff: The protection and deconfinement tradeoff (Tables 13-15 and Figure 2) serves as a cumulative indicator to evaluate the effectiveness of interventions, the balance between containment and relaxation, and wave behavior and dynamics.

Third, regarding intervention effect on containment: (1) Early containment: An epidemic would be quickly contained by implementing early and strong interventions at the every beginning of an outbreak when its cases are low. (2) Late containment: An epidemic would be contained by implementing strong interventions in a late stage of an outbreak when its case numbers are high. (3) Fast containment: An epidemic with high case numbers would be contained quickly by implementing strong interventions such as strict full lockdowns. (4) Slow containment: An epidemic with considerable case numbers would be contained slowly if weak interventions are implemented (probably also followed by strong interventions later) in the increase stage, or early and fast relaxation interventions are implemented in the decline stage when the cases are still high. (5) Effective containment: The effectiveness of individual interventions (including strong interventions) is sensitive to its implementation timing and the outbreak severity. Generally strong interventions may be much less effective in quickly containing an already explosive outbreak compared to early interventions in the initial stage of an outbreak with low cases. For early effective containment, strong interventions would be much more effective than many soft interventions. To contain an already severe epidemic, full lockdown together with control measures like business and school shutdowns, banning public activities and staying at home is the only effective option.

Fourth, regarding COVID-19 resurgence: (1) Wave differences: Wave differences in a country and between countries result from different strictness, timing and number of interventions; a normally-distributed case curve is likely associated with early strong interventions without significant early and fast relaxation of these interventions; late and soft interventions would result in an exponential distribution of an outbreak for a longer period of curve with higher peak cases; prematurely early and fast relaxation of interventions would extend the curve to a long tailed distribution. (2) Resurgence: Prematurely relaxing or lifting strong interventions, early and fast reopening, and more infectious virus mutants may likely expand infection channels and cases or even change the case movement direction from a downturn to a uprising trend. (3) Protection-deconfinement conflict: The protection-deconfinement conflict determines the non-normal curve distribution of cases. A resurgence may occur when the deconfinement rate is substantially higher than the protection rate; higher difference more rapid growth of cases. In contrast, when the protection rate is much higher than the deconfinement rate, the curve will be flattened.

Fifth, regarding the impact of vaccination, without considering vaccine-induced immunity: (1) Under weak intervention: While Israel and Japan adopted vaccination in the period of the recent resurgences, weak interventions and more relaxations still resulted in higher and more infections. The simulation results of different protection-deconfinement scenarios do not obviously distinguish themselves from those unvaccinated countries. (2) Under virus mutation: The simulation results of the infection impact by increasing mutant transmissibility at different levels do not show difference from those unvaccinated countries. More infectious virus with weak interventions would still result in high infections and could change the case movement trend from downward to upward.

Lastly, regarding the containment-relaxation tradeoff and business reopening: (1) Containment-relaxation tradeoff in the increase stage: It would be difficult to reach a balance between effective containment and socioeconomic recovery in the increase stage of an outbreak since either late and soft intervention or early and fast relaxation may cause an exponential growth of cases. A balance of “living with the virus” may be possible in the increase stage of an epidemic only when considerable infection cases and moderate socioeconomic activities are acceptable. (2) Containment-relaxation tradeoff in the decline stage: In reality, socioeconomic recovery should mainly be pursued in the decline stage of an outbreak. Similar balance and arrangements as in the above increase stage would be more feasible in the decline stage, while such early reopening would result in a long tailed decline trend of infections. (3) Full reopening: It would be safe to fully reopen businesses and societies when cases are extremely low and good hygiene and self protection remain.

The aforementioned specific and general findings provide a tool to appropriately manage control and relaxation interventions, and estimate the impact of interventions on cases and the resurgence behaviors and trends. They inform policy-makers in making tradeoffs between epidemic containment and socioeconomic recovery, differentiating relaxation and control interventions, determining the timing of interventions, and when ‘living with the virus’ may be possible even under vaccination.

## Methods

### Data and Processing

We carefully chose and process the data fitting this data-driven discovery research. (1) Fatal SARS-CoV-2 virus mutations: we chose the waves relating to the original virus mutants alpha, delta and lambda which were more infectious and deadly, causing wider and more phenomenal effects; (2) COVID-19 waves and resurgences: relating to these mutations spreading over 2020 and 2021; (3) Vaccination conditions: both with and without vaccination periods; (4) Representative countries with different ethnic and regional backgrounds: in Europe including Germany, France and Italy, Middle East Israel, and Asia Japan; (5) NPI data: intervention polices and events relating to the time periods of the waves and countries manually collected from Wikipedia sources, which are categorized into two types: C for control policies, measures and actions; and R for lifting or relaxing restrictions; (6) COVID-19 case data: including active, recovered and death case numbers corresponding to the above periods and countries. Accordingly, the resurgences cover the first waves between 1 Mar. and 31 May 2020 and the second waves between 1 Sept. to 1 Dec. 2020 for Germany, France and Italy; from 1 Sept. 2020 to 20 Nov. 2020 and between 22 Dec. 2020 and 27 Mar. 2021 for Israel; and between 1 Dec. 2020 and 10 Mar. 2021, and the second is between 20 Mar. 2021 and 20 Jun. 2021 for Japan. Two waves of each country and their corresponding case numbers, mutants, vaccination, and NPIs are aligned. We forecast the next 10 to 30 day resurgence trends of COVID-19 infections under different severity, number and timing of interventions, public activities, variants, and vaccination conditions. More details about the data and processing are in the supplementary materials.

### Models

We propose a dynamic intervention event-driven interactive epidemiological compartmental model iSPEIQRD. iSPEIQRD invovles multi-aspect factors: epidemiological attributes, NPI - event attributes, pharmaceutical intervention (PI) - virus mutation. It further models the influences and interactions between NPIs, vaccination, virus mutation, the corresponding population’s epidemiological states, and the transition between people’s states. The structure of iSPEIQRD is shown in Figure 6. Our model (1) characterizes the time-varying dynamic epidemic process and its epidemiological attributes of the COVID-19 pandemic, as by other time-dependent SEIR structures^11,41^, (2) quantifies the interactions and individual and cumulative impacts of control (*i*_*C*_) and relaxation (*i*_*R*_) events on people’s epidemiological states, (3) differentiates the epidemiological and event attributes over the first-to-second wave evolution, and (4) makes these attributes adjustable for what-if analysis and simulation in terms of various control-relaxation intervention strategies, various transmissibility levels of virus mutations, and their joint effects on wave dynamics and first-to-second wave transform as well as COVID-19 resurgence under unvaccinated and vaccinated conditions. These are critical to connect COVID-19 epidemic and its mathematical compartmental modeling to their external driving factors such as government and public responses. iSPEIQRD further characterizes how interventions affect the dynamic transfer from the first to second waves, and how control or relaxation interventions affect protection and deconfinement and eventually the epidemic dynamics individually and cumulatively in complex conditions such as with contrasting intervention strategies and more infectious virus mutations.

**Figure 6.**
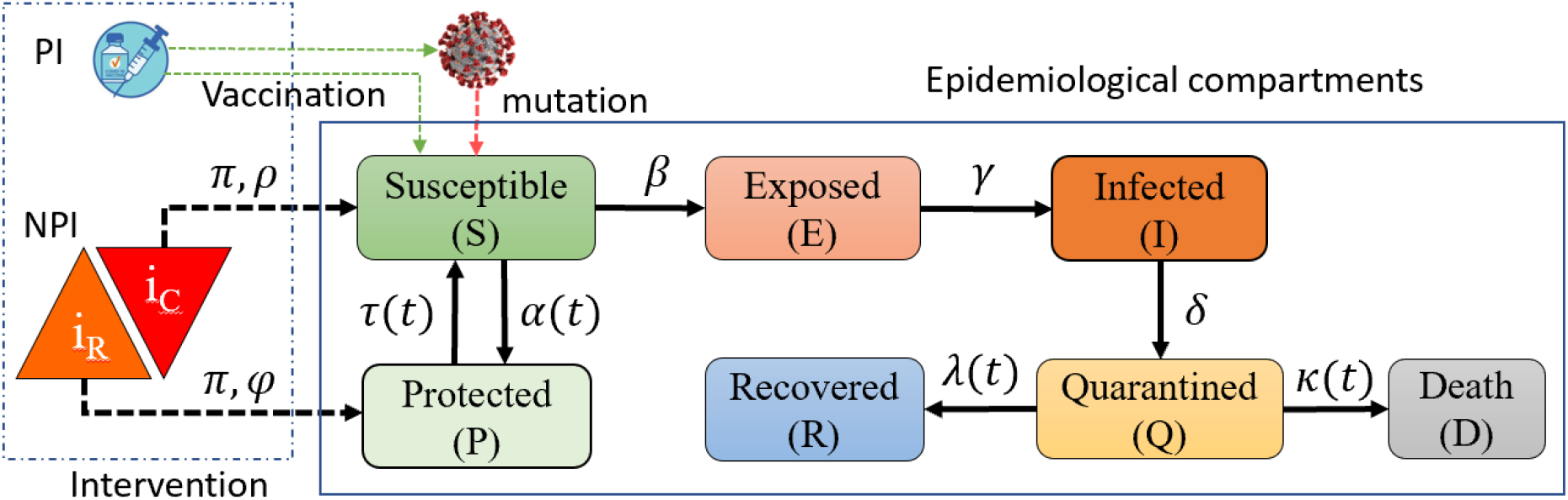
The dynamic intervention-driven compartmental model iSPEIQRD for modeling the interactions between interventions and epidemiological compartments in terms of their respective attributes and impact on people’s epidemiological state transition in a COVID-19 wave. Intervention impact attributes: the intervention impact *π*, the control indicator *ρ*, the relaxation indicator *φ*, the protection rate *α*, and the deconfinement rate *τ*; and epidemiological attributes: the infection rate *β*, the incubation rate *γ*, the quarantine rate *δ*, the recovery rate *λ*, the mortality rate *κ*, and the virus mutation on *β*. Intervention events and *α, τ, λ* and *κ* are time-varying.

First, during the pandemic evolution, governments sequentially enforce control measures to flatten the curves during outbreaks (i.e., control interventions *i*_*C*_ in Figure 6) or relax restrictions (i.e., *i*_*R*_) to restore socioeconomic activities when infections substantially decline or diminish. To model such different stages and strategies of enforcing or relaxing interventions and their effects, iSPEIQRD incorporates the interactions between these control and relaxation interventions and epidemiological states into the epidemic transmission processes and models their influences on cases and state transition by an event-driven dynamic interactive model. An *event* refers to either an intervention that is undertaken to control the COVID-19 pandemic (or unwind it) or a public activity such as a protest or a public gathering that has a great influence on the trend of COVID-19 cases. Specifically, *control interventions* (C) are the COVID-19 countermeasures implemented during infection spread and outbreak; *relaxation events* (R) are policies and activities implemented during the social recovery period to relax or unwind the controls. Besides government interventions, public activities such as large-scale gatherings happening during outbreaks may also influence the COVID-19 curves^42^. For example, in Germany, two large protests on 1 Aug. and 20 Sept. 2020 in which a majority of the participants ignored the mask-wearing and social-distancing requirements likely contributed to the dramatic increase of COVID-19 cases after a few days. Such maskless gatherings could be an important contributor of COVID-19 resurgence even though it is challenging to infer their specific influence directly. We thus involve them as relaxation events.

Second, as an epidemiological model, iSPEIQRD partitions the entire population into seven epidemiological compartments, as shown in Figure 6, to characterize seven unique states of population involved in an epidemic and their state transition over evolving COVID-19 epidemic. The population flow during the epidemic transmission at time *t* is characterized by Equations (1)-(7), where *S* is the susceptible population, *E* is the exposed population, *I* is the actual infected case number which contains unreported cases, *Q* is the quarantined case number, *R* is the recovered number, *D* is the death number, *P* is the protected and confined population, and *N* is the total population. The protected or confined population in the compartment *P* refers to those who are not susceptible to the disease due to their good self-protection habits or staying away from infectious people. Additionally, we suppose *N* is a closed population and ignore the natural birth and death numbers as they are negligible compared to the large target population in *N* and the short time period of each wave.

iSPEIQRD involves two sets of parameters. (1) Intervention attributes: the indicators *ρ* for control events and *φ* for relaxation events, the protection rate *α* reflecting the influence of control events on enforcing containment, the deconfinement rate *τ* reflecting the influence of relaxation events on relaxing containment, and the change of infection *β* caused by mutation. (2) Epidemiological attributes: the infection rate *β*, the incubation rate *γ*, the recovery rate *λ*, the mortality rate *κ*, and the quarantine rate *δ* measuring the quarantine effect on restricting infected people in a quarantined state. The parameters *α, τ* and *δ* characterize the influence of control and relaxation events and quarantine on the epidemic. A positive protection rate *α* models the improvement of public health and self-protection awareness, such as encouraging wearing face masks and good personal hygiene (e.g., hand sanitization). Similarly, a positive deconfinement rate *τ* simulates the return of those people from the confined compartment to the susceptible compartment due to their potential loss of acquired immunity, the deconfinement measures (e.g., society reopening), or other unprotected activities (e.g., large-scale gatherings without mask wearing).

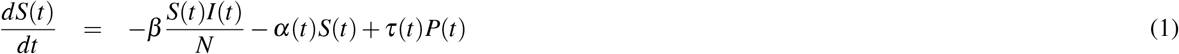

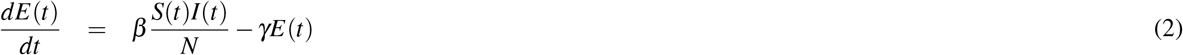

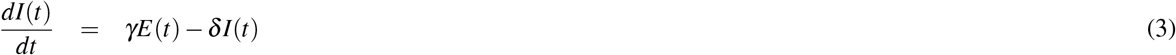

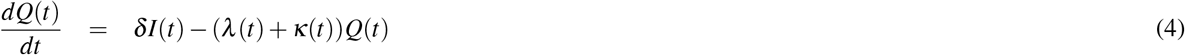

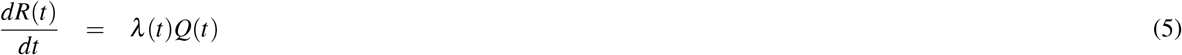

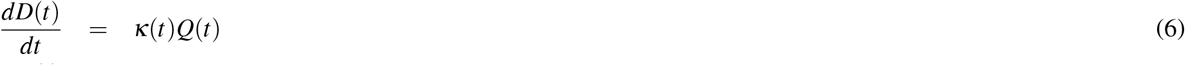

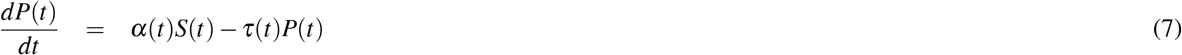

To quantify the event effects on COVID-19 transmissions, both the protection rate *α* and the deconfinement rate *τ* are assumed to be event-driven and follow step functions shown in Equations (8)-(9), where *m* refers to the total number of events, and *ρ* and *φ* are indicators for control events (i.e., type C in Tables 3-5) and relaxation events (i.e., type R in Tables 3-5), respectively. *π* represents the impact level of a control or relaxation intervention (or a public activity). Specifically, *ρ*_*n,t*_ equals 1 if the *n*^*th*^ event is a control measure and in place at time *t*; otherwise it equals 0. Similarly, *φ*_*n,t*_ equals 1 if the *n*^*th*^ event is a relaxation policy and in place at time *t*; otherwise it equals 0. When control measures are ongoing, more people will be protected from being infected under the stricter measures, and their self-protection awareness is also enhanced, correspondingly increasing the protection rate *α*, as shown in Equation 8. However, to revive social and economic activities, governments may relax restrictions progressively, resulting in more people moving back to the susceptible compartment since the virus is still valid, i.e., increasing the deconfinement rate *τ* as shown in Equation 9. Large-scale public activities such as protests may violate control measures to convert some protected individuals to susceptible, which is also shown in our design and experiments. Since there is a time delay between an event being undertaken and its actual effect being validated, we introduce the time delay parameter *ν* as an integer on a daily basis to the step functions and infer its value by fitting the case data. The time delay parameter *ν* reflects the *ν*-day delayed effect of intervention *n* implemented on day *t* on enforcing or relaxing containment.

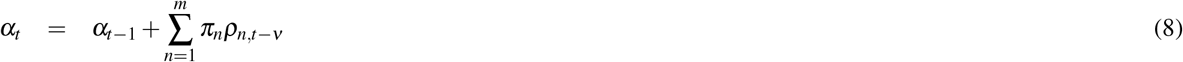

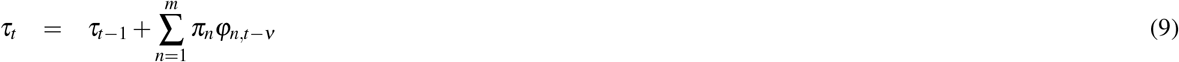

Further, epidemic dynamics and the dynamic interactions between interventions and compartmental transition are captured by the time-varying parameters: the recovery rate *λ*, the mortality rate *κ*, the protection rate *α*, and the deconfinement rate *τ*. These parameters measure both dynamic states of populations in the epidemic transmission and the sequential control and relaxation interventions and their evolving impact on the epidemic transformation over time. This captures the interactions between epidemiological attributes and intervention impact attributes. As shown by the case movements in Figures (8)-(10), the recovered and death populations evolve over time, i.e., described by the time-dependent cure and mortality rates as explored in^11,41,43^. The estimated recovered and death rates are not constant in both the first (which are complete) and second (which are partial and still evolve at that time) waves in Germany, France and Italy, except the recovery rate of the second wave in France. To capture the different trends of recovered and death cases affected by the interventions, quarantine and protection, we simulate the recovery rate *λ* and the mortality rate *κ* in terms of three optional exponential functions respectively in Equations (10)-(11).

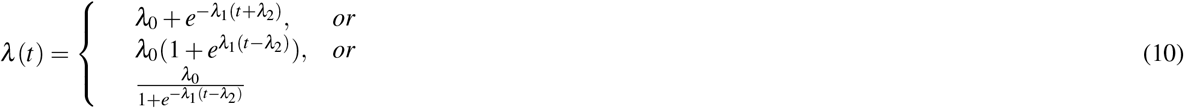

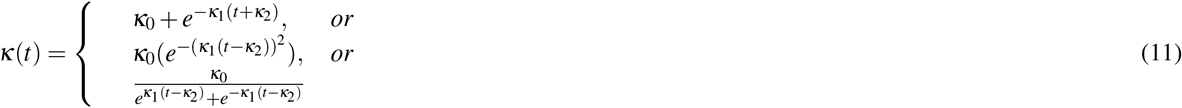

Specifically, unlike studies that treat the infection rate as time-dependent^44,45^, we consider it constant since this may better capture the intrinsic epidemiological attribute of the virus in a fixed short period of a wave, when the infection ability of the virus likely remains stable as interventions are enforced to protect people and contain transmission. On the other hand, the dynamics of infection rate observable from the daily cases is sensitive to the enforcement and adjustment of control or relaxation policies and people’s increasing self-protection awareness and response to the epidemic. To capture the influence of these factors on infection, we introduce the protected compartment for those people under protection during transmission and further measure the influence of enforcing or relaxing interventions on these people in terms of dynamic protection and deconfinement rates. In addition, to characterize the virus mutation, we adjust the infection rate as more infectious virus mutants would incur higher transimissibility.

We solve the model with a nonlinear data-fitting approach that minimizes a least squares error function as shown in Equation (12), where *F* represents our model, **x** denotes the input data (*Q, R* and *D*), provided by the integration of the ordinary differential equation (ODE) system (Equations (1)-(7)) and solved with the fourth-order Runge-Kutta method. The Runge-Kutta method is easy to implement and very stable^11^. *y* denotes an observation (i.e., the reported active case number, the recovered case number, and the death case number). *θ* refers to all parameters (*α, β, γ, δ, κ*_0_, *κ*_1_, *λ*_0_ and *λ*_1_) that are inferred by Equations (1)-(11).

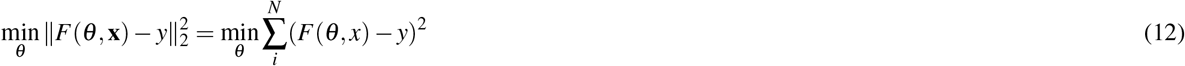

This function requires initial values for optimization. We set the initial values {0.1, 2.0, 0.2, 0.1, 0.1, 3} for the unknown parameters {*α, β, γ, δ, τ, ν*}, respectively. Some parameters are fine-tuned during the training process for each country. The optimization problem described in Equation (12) is subject to the constraints specified by the lower bound *l*_*b*_ = [0, 0, 0, 0, 0, 0] and the upper bound *u*_*b*_ = [1.0, 5.0, 1.0, 1.0, 1.0, 10]. The initial values of the event impact are all set as 0.001 with the lower bound 0 and the upper bound 1. In addition, the initial conditions of the exposed, infected and protected cases *E*_0_, *I*_0_, *P*_0_ are unknown in the public dataset, which are unlikely to be zero. Hence, we set them as 0.1*Q*_0_ since our model is not sensitive to these initial conditions. The representative measure of the optimal set of parameters is obtained with up to 2,000 iterations of optimization under the initial values and the constraint of the bounds.

## Data Availability

The source codes and data are shared in Github, with the hyperlink included in the paper.

https://github.com/QingLIU67/waves_comparison

## Additional information

## Acknowledgements

Thanks to Hu Cao for contributing to some data collection and experiments. Thanks those readers with comments on the preprint version shared in medRxiv.

## Author contributions

LBC contributed to conceptualization, methodology, modeling, and writing. QL contributed to modeling, experiments, and editing.

## Data and materials availability

The data and codes of the original version are available at GitHub: https://github.com/QingLIU67/waves_comparison.

## Supplementary materials

### Data and Processing

For countries without vaccination and virus mutation influence but had clear wave differences, we choose the first and second waves in Germany, France and Italy in 2020 for case studies. For vaccinated countries affected by virus mutations such as delta, we choose the two recent resurgences in Israel and Japan. These five countries are located in Europe, Middle East and Asia, representing different ethnic and cultural backgrounds, having different intervention strategies and vaccination conditions, and spreading over 2020 and 2021 (^5^, the variants from https://covariants.org/per-variant, and the vaccination data from https://ourworldindata.org/covid-vaccinations.). Table 1 summarizes the start and end dates of two waves, 10-day prediction, and 30-day prediction. Table 2 shows the dates when the first alpha or delta virus strain was identified and the first and second dose vaccination was undertaken in each country. In Germany, France and Italy, their first waves are specified as between 1 Mar. and 31 May 2020 and the second waves between 1 Sept. to 1 Dec. 2020. Each wave has 92 days for the alignment and comparison between two waves vertically and between countries horizontally. These two waves in each country clearly showed the first-to-second wave evolution with clear wave differences and different intervention strategies. The two resurgences in Israel spread from 1 Sept. 2020 to 20 Nov. 2020 and between 22 Dec. 2020 and 27 Mar. 2021, respectively. The first wave was assumed unaffected by virus mutation and vaccination, while the first virus mutant alpha was identified on 14 Dec. 2020 at the end of the first resurgence, the first dose vaccination started on 20 Dec. 2020, and the first delta infection was detected on 5 Apr. 2021 outside the second resurgence. In Japan, the first resurgence corresponds to the period between 1 Dec. 2020 and 10 Mar. 2021, and the second is between 20 Mar. 2021 and 20 Jun. 2021. The first alpha infection was pronounced on 18 Jan. 2021 in the first resurgence, the first dose vaccination started on 10 Jan. 2021, and the first delta infection was identified on 10 May 2021 within the second resurgence.

We collect the ground truth information of daily cases and sequential interventions and public activities aligned with the above two waves in each country and analyze the influence of interventions and virus mutation on cases and explain our modeling objectives and findings. We use the JHU CSSE COVID-19 data^46^, which records the worldwide daily case reports including total confirmed cases, recovered cases, and deaths. The intervention event information is collected from Wikipedia for the COVID-19 pandemic in Germany^6^, France^7^, Italy^8^, Israel^9^, and Japan^10^. It includes both government-initiated mitigation policies and public activities, which influence COVID-19 transmission and resurgence. The OxCGRT (the Oxford COVID-19 Government Response Tracker) also collects systematic information on government responses to COVID-19 across countries in four categories: containment and closure, economic response, health systems, and miscellaneous^47^. However, the policy information recorded in this dataset is somewhat inconsistent and less timely than the events reported above. Moreover, large public events that may influence the COVID-19 trend in a country are excluded from OxCGRT. We therefore collect event information from the Wikipedia sources.

In this analysis, we only include the events that happened during the periods of the first and later waves or resurgences for the alignment between epidemiological attributes and intervention impact attributes. In addition, the cases recorded between the end of the first wave and the beginning of the resurgence are stable and the released policies show less significant impact on the more stabilized epidemic curves. Therefore, we exclude such periods in the two waves for all countries. As shown in Tables 3-7 for Germany, France, Italy, Israel and Japan, the extracted major events are classified into two types: *C* for control policies, measures and actions; and *R* for lifting or relaxing restrictions. During the two time periods, the target countries experienced different first and second/later waves or resurgences of COVID-19. Their governments implemented different control and relaxation policies in response to the two outbreaks over different stages of the waves and their case movements. Their policies changed from the initial control-oriented measures in the first wave to relaxation-favored strategies after the first wave was contained and then to control measures again to contain their COVID-19 resurgence.

### Additional results

The existing studies mainly focus on specific aspects and a qualitative to descriptive analysis of second waves and resurgences. The contrasting epidemiological characteristics between waves remain largely unknown. The epidemiological predictive analysis of multi-waves has received limited attention, leaving significant gaps in areas including deep epidemiological characterization and simulation of the resurgence and deep insight into how various interventions affect the resurgence. In reality, the later waves become more global, repetitive, infectious, and variable, particularly with the evolving mutants. There are many important questions to be answered by evidence. For example, what leads to the second waves? Why are later waves more severe (e.g., in terms of cases and impacts) than the first? Is the knowledge learned from the first wave transferable to better control later waves? How does virus mutation affect the epidemic? In addition to the above results, here we present extra results which together offer comprehensive evidence and insights about the dynamics, evolution, attributes and causes of COVID-19 resurgence and the influence of external events and virus mutations on the resurgence for people with and without vaccination.

### What do we know about COVID-19 resurgence?

The resurgence over multiple waves can be illustrated by what happened in some European countries. At the beginning of the first wave in early 2020, most European countries adopted and adjusted a series of strong interventions, which successfully reduced cases and the trajectory of growth. Taking Germany as an example, their cases almost doubled within one day on 1 March 2020. Various interventions were then implemented, including but not limited to cancelling large-scale events, shutting down schools, childcare and businesses, and enforcing restrictions on gathering. Their step-wise increase of these control measures flattened their case curve two months later. However, just a few months later, many European countries had resurgence even worse than their first waves after prematurely or too quickly relaxing their interventions and reopening borders and businesses. As shown in the daily case reports (Figure 7), their second wave peaks were significantly higher than their first waves. For example, in Italy, the peak daily cases reached 6,554 in the first wave, compared with 40,896 cases on 13 November in the second wave, over six times higher than the first peak. Similar differences between waves appeared in most European countries in 2020 and in many other parts of the world in 2021, such as Australia, Vietnam, South Korea, and the USA.

**Figure 7.**
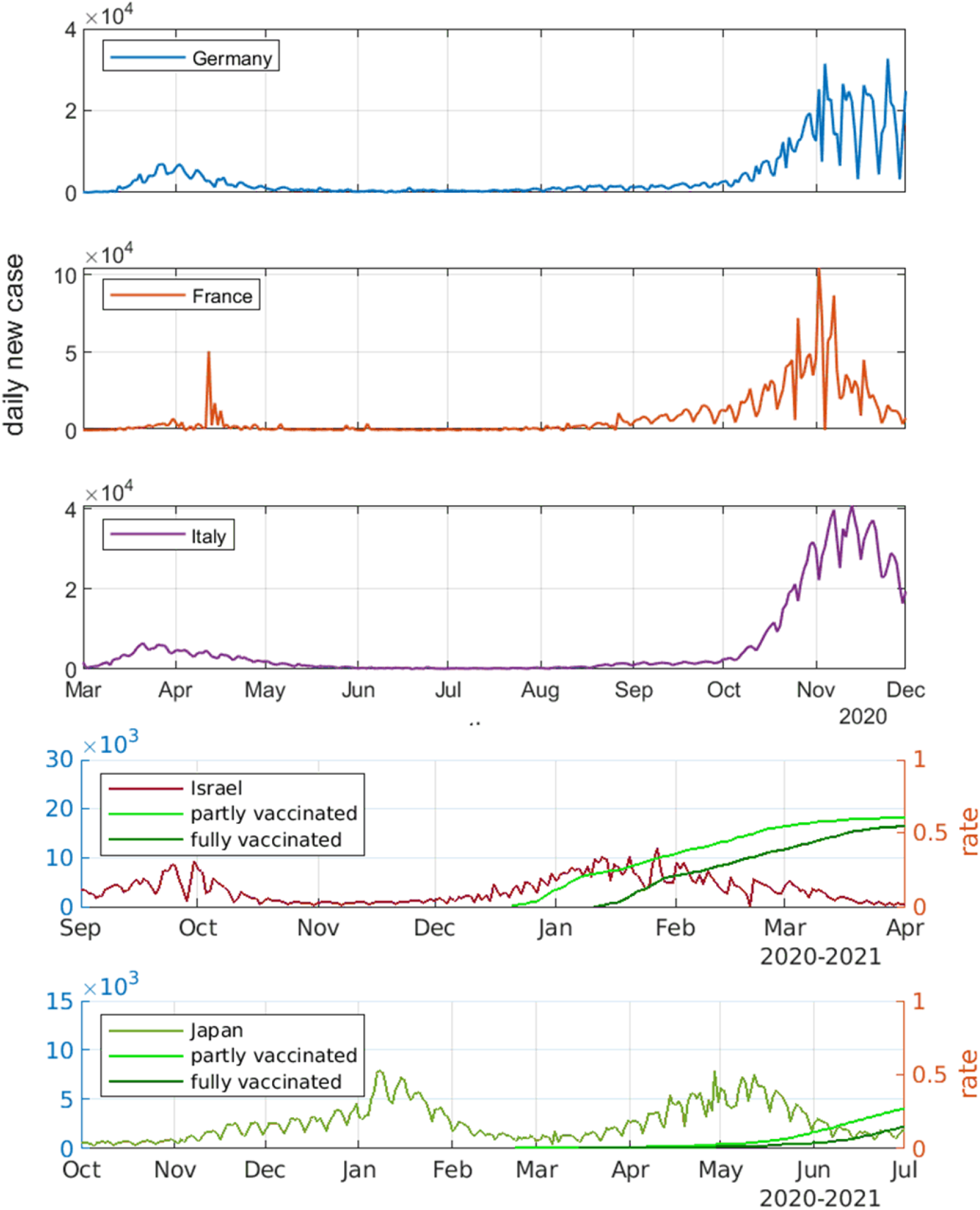
Daily new cases and first-second dose vaccination rates in Germany, France and Italy from Mar. to Dec. 2020, in Israel from Sept. 2020 to Apr. 2021, and in Japan from Oct. 2020 to Jul. 2021. Two evolving waves of COVID-19 epidemic appear in each country, with the five countries (1) having respective resurgence attributes and wave patterns; (2) vaccination in Israel (started on 20 Dec. 2020) and Japan (10 Jan. 2021); (3) alpha and delta strains detected in Israel (alpha on 14 Dec. 2020, delta on 5 Apr. 2021) and Japan (alpha on 18 Jan. 2021 and delta on 10 May 2021). The left y-axis refers to daily new cases, and the right y-axis refers to first-second dose vaccination rates.

However, as shown in Figure 7, Israel and Japan show different wave evolution and patterns from the three European countries. Though their second resurgences involved more relaxed interventions, both waves are associated with more similar case movement patterns including similar peak case value and case distributions than Germany, France and Italy. The lower peak cases and daily cases under soft containment in Israel may have benefited from their early vaccination and high vaccination rate. In Japan, though the vaccination started at the beginning of their second resurgence, the vaccinate rate was very low over most of the second wave period. The overall daily cases are much higher and denser than their first wave when stronger interventions were implemented, which may be related to the fact that both alpha and delta strains were identified in Japan over the second wave.

There may be various significant and distinct factors driving the COVID-19 resurgence and their differences from the first waves in individual regions, including the more infectious viral mutants. A critical and common trigger is the premature and fast relaxation of interventions, including hygiene behaviors, social and business restrictions, and border control. Despite the positive effect of interventions on flattening the curves, the harsh interventions, and the ‘new normal’ also had significant negative effects on individuals, society, and the economy. Enforcing strong interventions such as social distancing and gathering restrictions significantly inconveniences people’s daily life. Travel bans, lockdowns, and curfews also seriously disrupt social and economic activities, increasing unemployment, poverty and inequality with adverse psychological impacts and pressure on governments. Governments face the dual burden of both providing financial support and being pressured to reopen and recover^4^. Consequently, after months of strict interventions, requests or demands for increasing psychological, behavioral, public, and political relaxation emerge over time and spread from one region to another. Subsequently, most countries relaxed their interventions by easing or lifting social distancing restrictions, and reopening schools and workplaces. However, the virus has not been eliminated and even worse mutants have emerged and spread through households, strangers, communities or airborne transmission. The fast relaxation of interventions supports virus transmission. This eventually causes multiple waves and resurgence.

### How do epidemiological attributes change over the different waves in different countries?

The epidemiological attributes of the two waves in Germany, France, and Italy without virus mutations and vaccination in 2020 are inferred by the iSPEIQRD model, shown in Tables 8-10, respectively. In contrast, Tables 8-10 show the epidemiological attributes of two resurgences over 2020 and 2021 with virus mutations and partial vaccination in Israel and Japan. These two groups share both similar and contrasting results. In the three European countries, first, the infection rates of the first waves are much higher than those of the second waves, showing a higher infection probability of the susceptible individuals in the first waves, which may have lifted people’s self-protection awareness in the second waves and resulted in lower infection rates. Second, the reciprocal of the incubation rate *γ*, namely *γ*^*−*1^, reflects the average incubation period. The shorter incubation rates in the three countries indicate longer incubation periods in their second waves. For example, France had an average incubation period of 2.2 days in the first wave compared to 6.6 days in the second wave, inferred from their case data. The longer incubation periods may also involve more cases infected in the second waves. Third, the quarantine rates *δ* increased from the first waves to the second waves in both Germany and France. The exception in Italy, however, cannot be simply explained as that was caused by fewer people quarantined in the second waves. It is possible that, due to the stretched healthcare resources, many more identified infections in the second wave may have caused a relatively lower quarantined ratio.

In the second group, the similar epidemiological attributes of the two waves to Germany and France are seen in Japan, as shown in Table 12. This may be owing to the similar mitigation strategies and people’s response, although the virus mutants alpha and delta were identified in the two waves respectively and the vaccination rate was low thus played a minor role in preventing infection. In contrast, the two waves in Israel show results contrasting to the other four countries, as shown in Table 11. Their infection rate, incubation rate, and quarantine rate of the second resurgence are all higher (30% to 60% increase) than that of the first wave, although the vaccination program started in the early stage of the second wave and the vaccination coverage was substantial and the highly infectious delta was not detected then. The incubation period reduced from 4.44 days in the first wave to averagely 2.74 days in the second resurgence, which may be related to its 32.6% higher quarantine rate. However, the infection rate in the second wave is 35.8% higher, this may be related to the spread of the alpha strain and relatively weaker and more relaxed interventions and an early easing of lockdown.

Further, we analyze the characteristics of recovery rate and mortality rate of two waves in the five countries. Figures 8-12 show the actual and inferred rates of two waves in each country. Figures 13 and 14 further compare the estimated recovery and mortality rates in two waves over five countries, estimated by iSPEIQRD, which clearly show the relations and differences between countries in the first and second waves. Below, we further discuss the findings.

**Figure 8.**
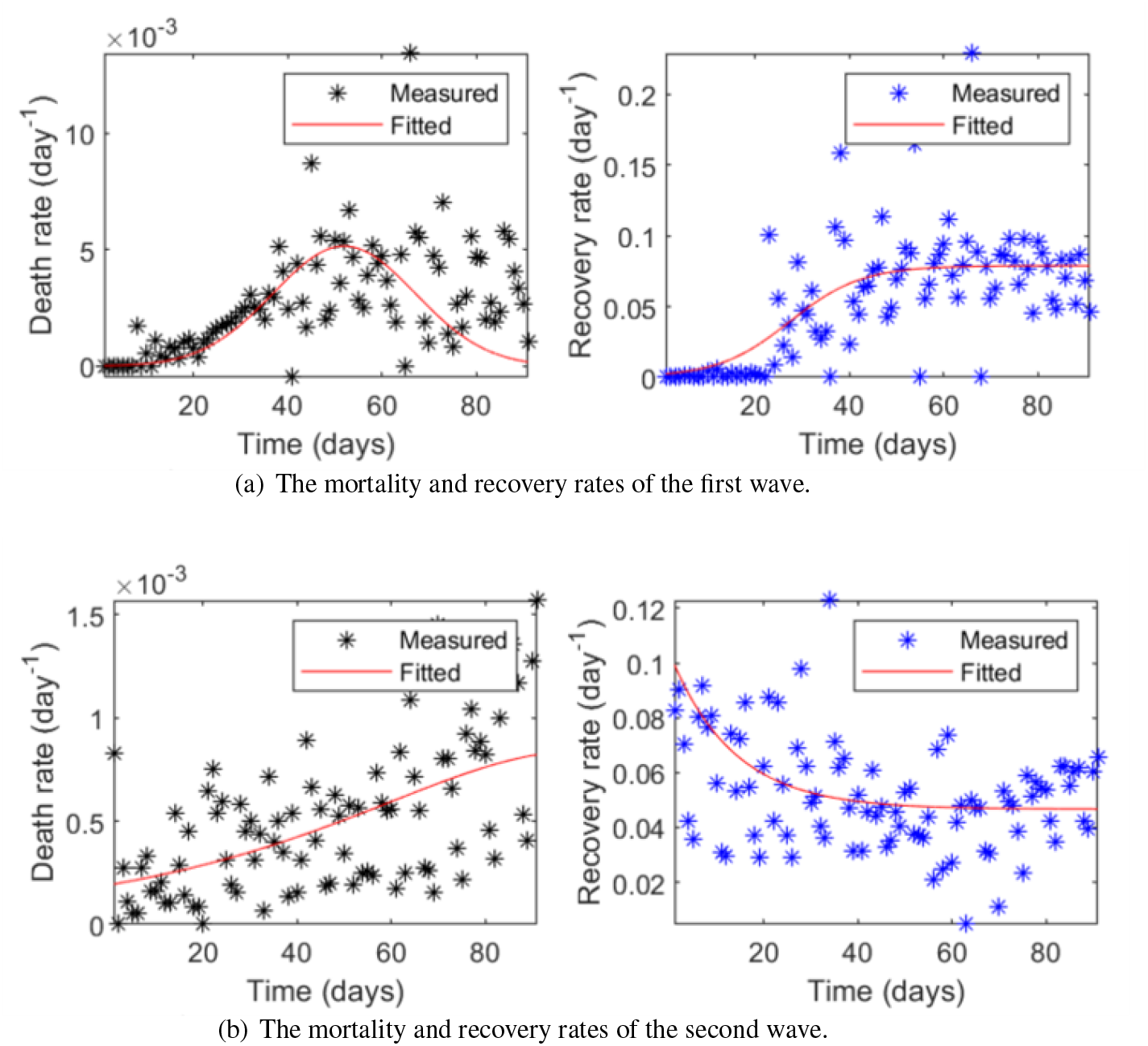
The time-varying recovery and mortality rates of two waves in Germany.

**Figure 9.**
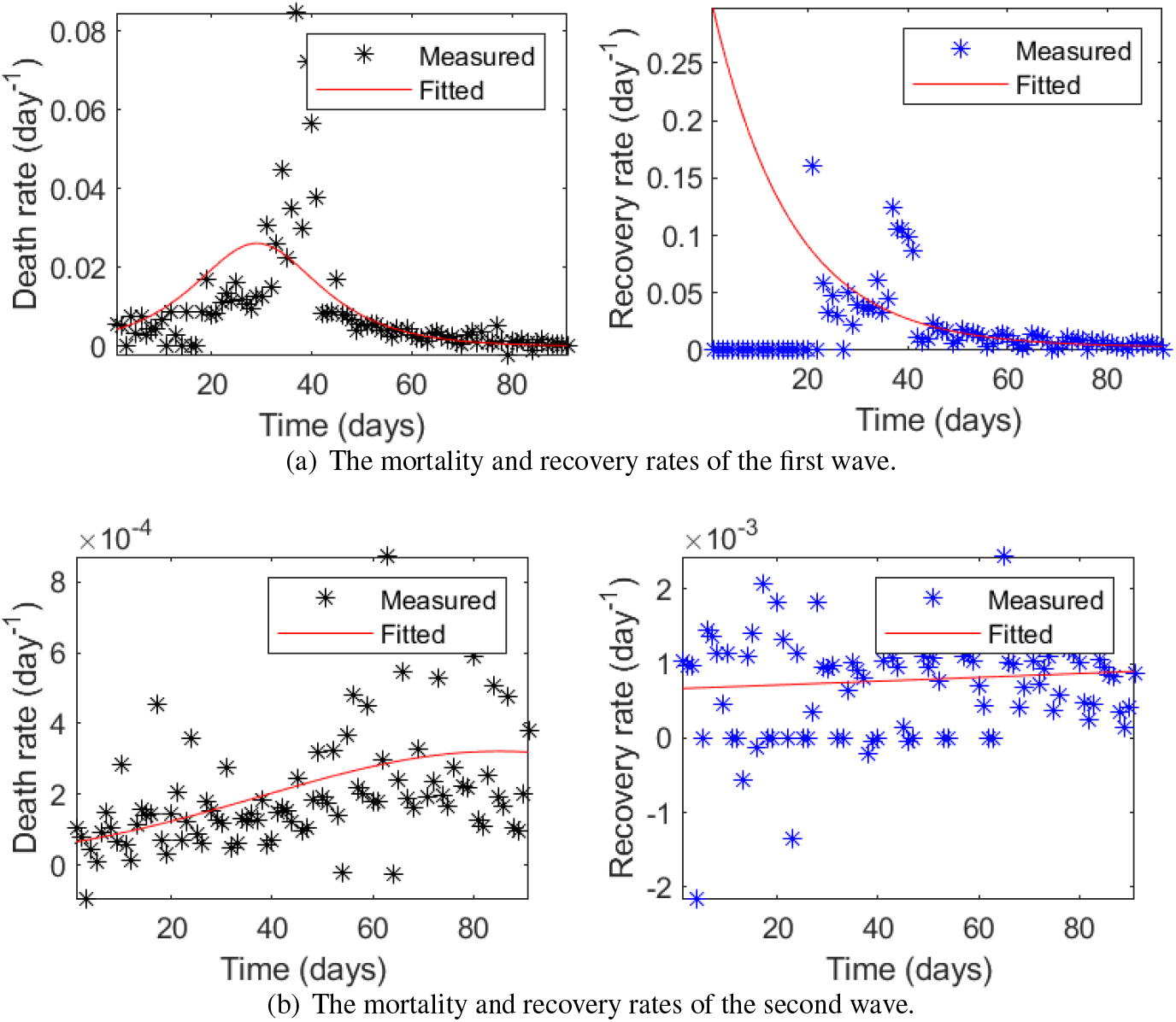
The time-varying recovery and mortality rates of two waves in France.

**Figure 10.**
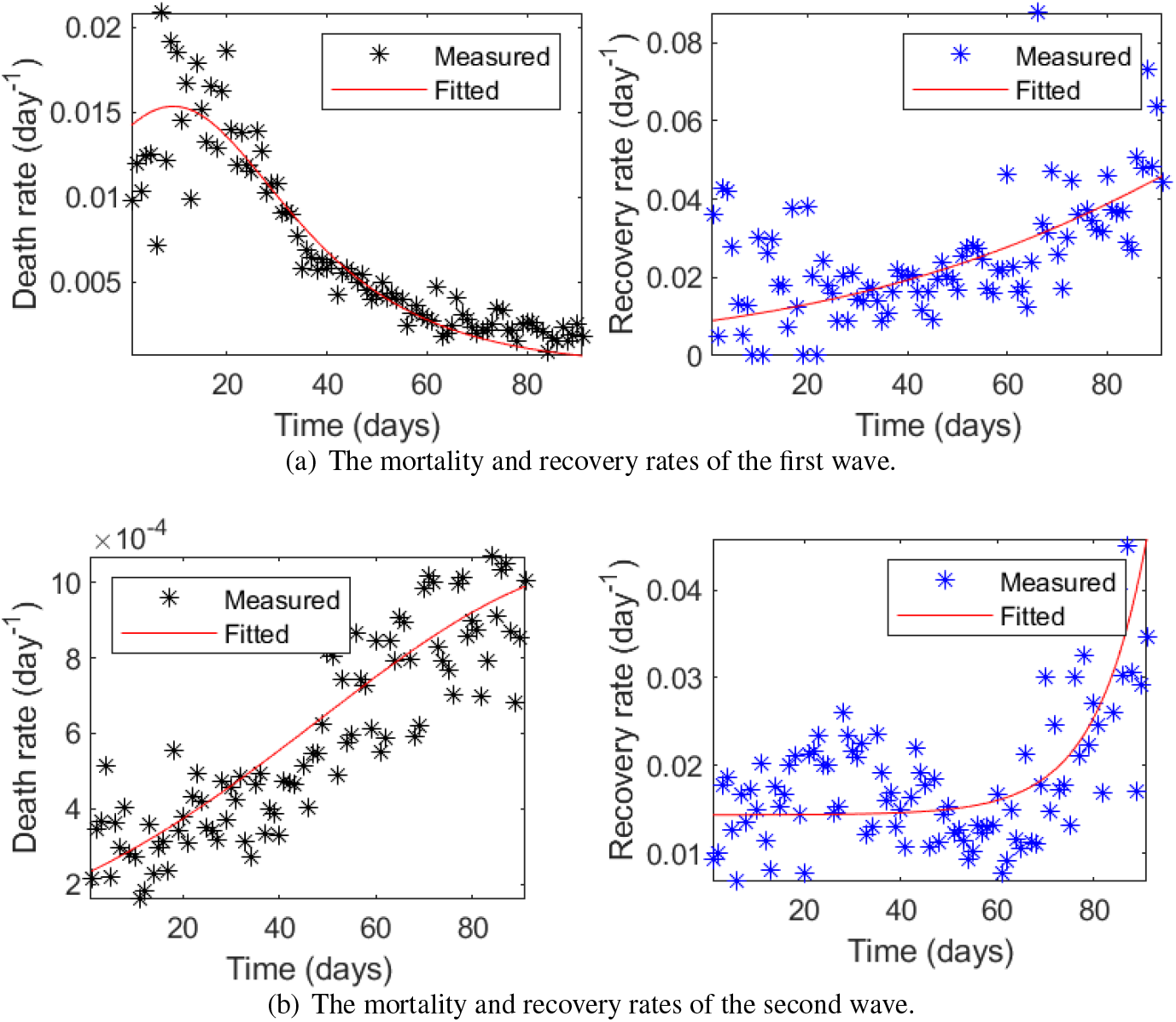
The time-varying recovery and mortality rates of two waves in Italy.

**Figure 11.**
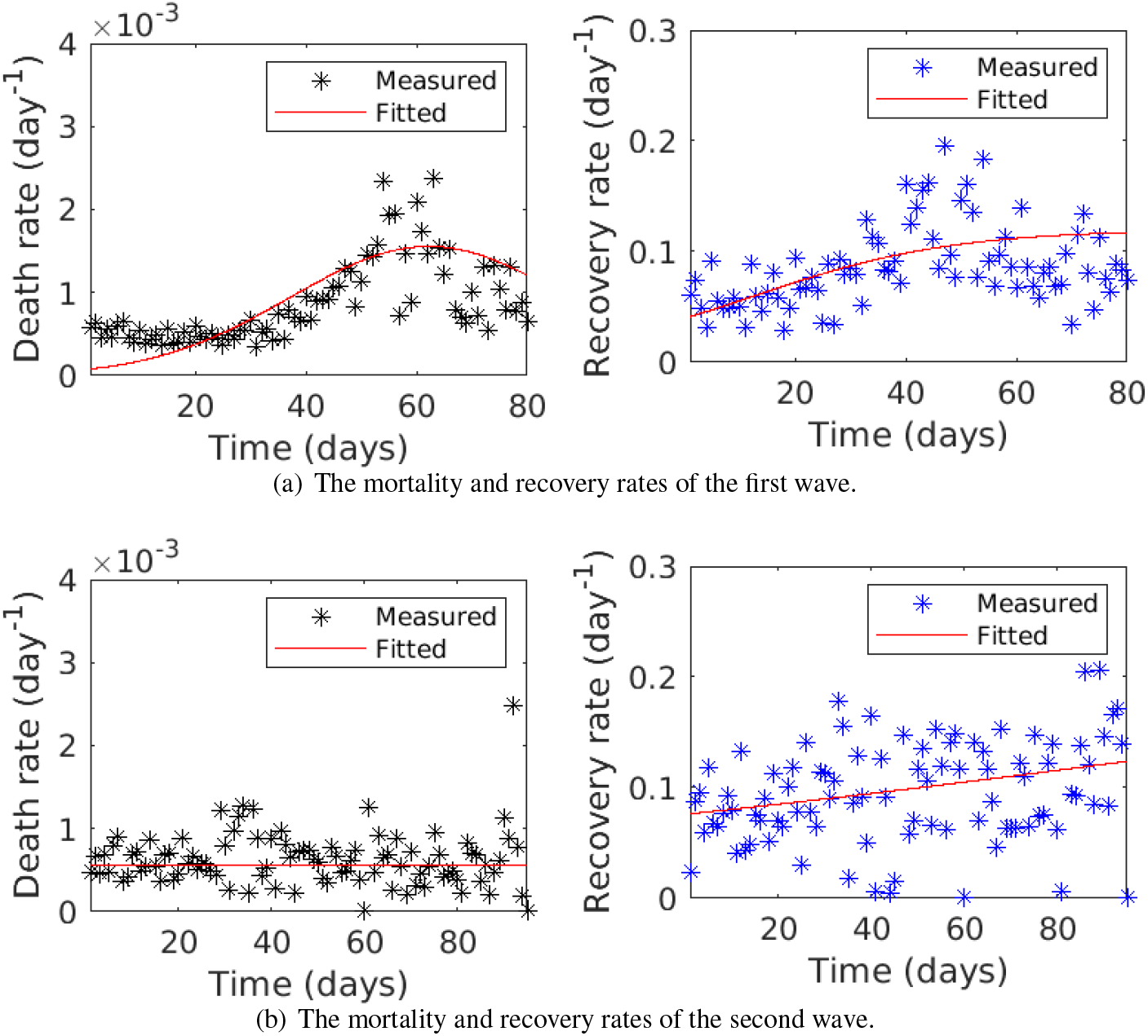
The time-varying recovery and mortality rates of two waves in Israel.

**Figure 12.**
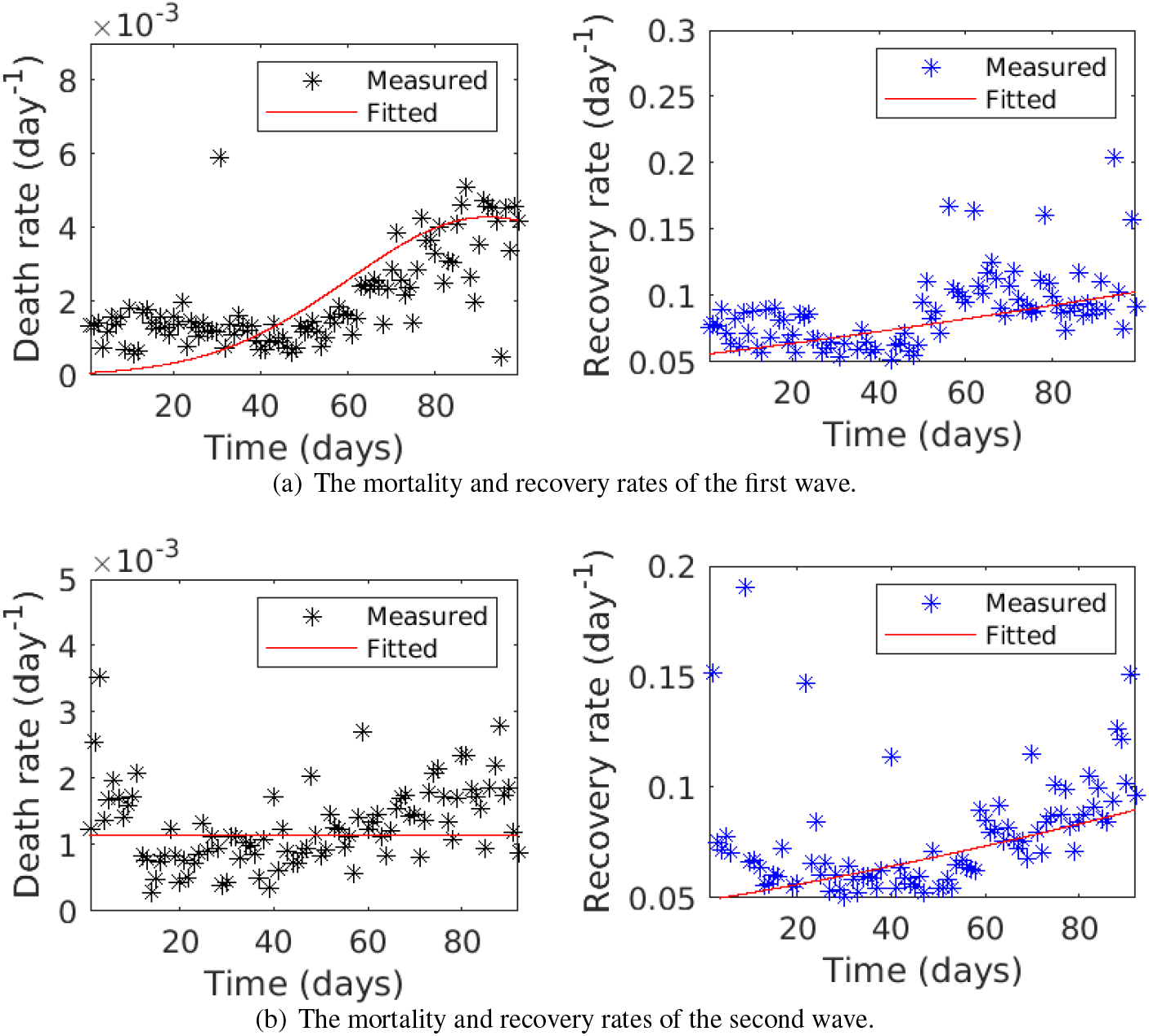
The time-varying recovery and mortality rates of two waves in Japan.

**Figure 13.**
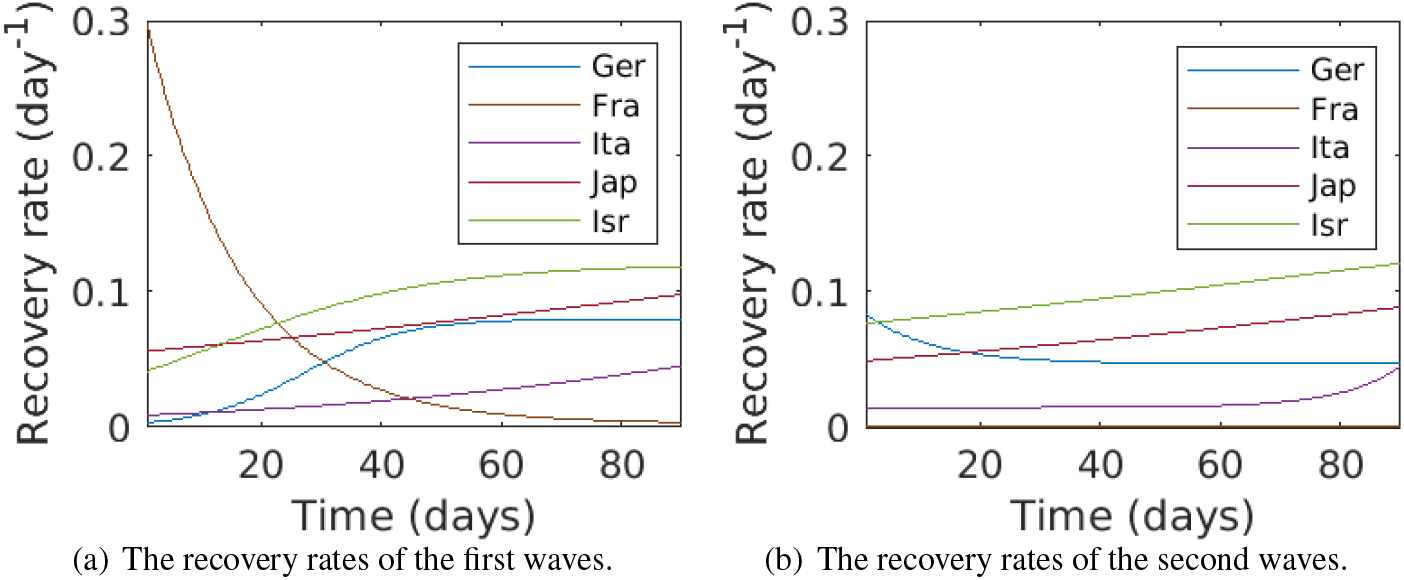
The estimated time-varying recovery rate of two waves in the five countries by the iSPEIQRD model.

**Figure 14.**
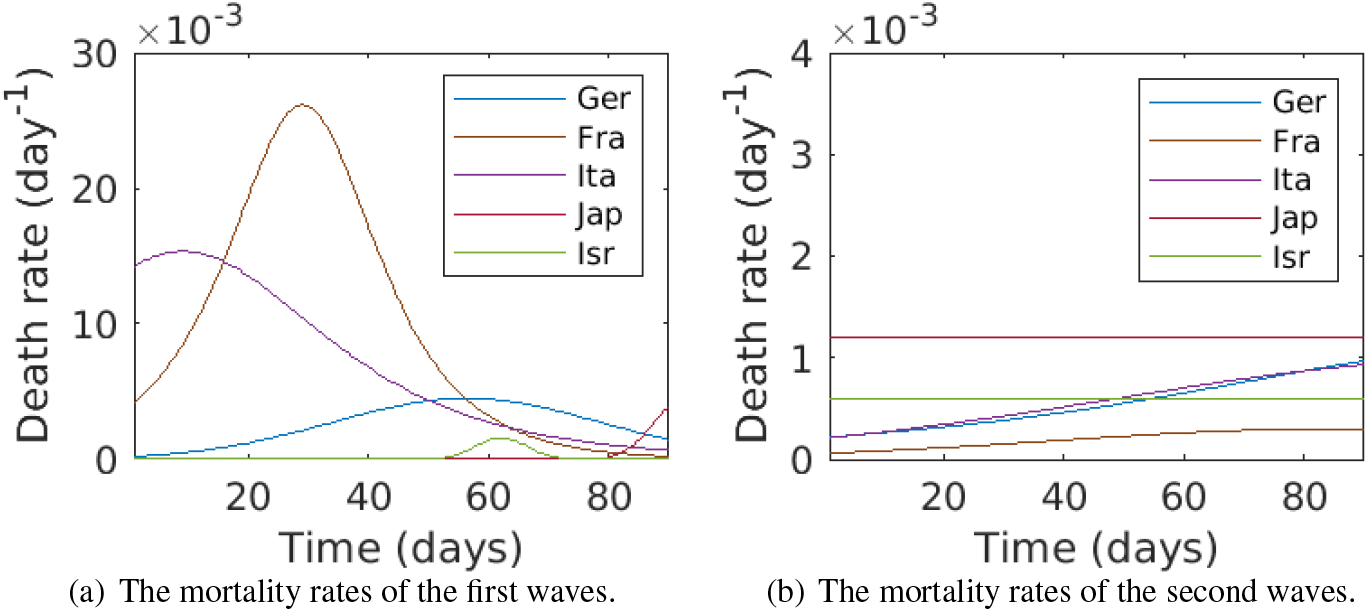
The estimated time-varying mortality rate of two waves in the five countries by the iSPEIQRD model.

By assuming that both the recovery rate *λ* and the mortality rate *κ* are time-varying, Figures 8-10 show the time-dependent movements of the recovery and mortality rates inferred from the case data in Germany, France, and Italy. In reality, the measured recovery and death rates are not constant in both the first waves which were complete and the second waves which were still evolving during that time period, although the recovery rate of the second wave in France tends to be more stable. The three countries share similar mortality changes in both waves. Specifically, in the first waves, their death rates decreased to stable after their initial increase, while they all rose gradually in the second waves. The recovery rates were more variable than the mortality rates, which stabilized at a certain level in Germany and France, in contrast to Italy. Taking Germany as an example, as shown in Figure 8(a), the death rate increased until it reached the peak at around 0.005 and then gradually decreased to a low level in the first wave. The recovery rate followed the same increasing trend at the beginning but then stabilized at nearly 0.08 one month later. This phenomenon is likely attributed to the greater investment in medical resources such as PPE, ICU beds, and ventilators to contain COVID-19. However, Germany’s second wave shows a different trend in Figure 8(b). The death rate gradually increased and the recovery rate decreased until it reached a stable level lower than that of the first wave. There are two possible explanations: first, the greater number of cases in the second wave overwhelmed healthcare systems; second, unlike the first wave which is complete, the second wave was still underway thus the epidemiological attributes did not stabilize yet. In contrast, Israel and Japan share the similar trends of mortality and recovery rates of the two waves, as shown in Figure 11 and Figure 12, respectively. In Israel, the recovery rates of the two waves showed a similar trend, while the second wave seemed to have a higher recovery rate. The death rate of the first wave increased over time significantly, in contrast to a relatively flat and much lower rate of the second resurgence. The overall recovery and death rate of both waves were much lower than that in the three European countries. These could be highly related to the stricter interventions and greater activity restrictions as well as early and substantial vaccinations in Israel, although alpha strains spread over the second wave. The Israeli data shows, although the infection rate was higher in the second wave, the death rate was actually much lower and the recovery rate was much higher. This may be related to their substantial vaccination covering the entire second wave and the higher quarantine rate. In Japan, both waves hold very the similar trends of recovery and death rates to Israel. However, the recovery rates of both waves in Japan were lower than that of Israel, and the death rates of both waves in Japan were higher than that of Israel. This discrepancy between Israel and Japan may be related to (1) higher overall control in both waves, and (2) earlier vaccination and higher vaccination rate in Israel than in Japan, as shown in Table 16 for Israel and Table 17 for Japan. This difference may also explain that vaccination is helpful for containing death and improving recovery.

In addition, though the epidemiological attributes are subject to other driving factors beyond interventions, we also obtain the following general epidemiological features of the second waves in the five countries, evidenced by the results in Tables 8-12: (1) a lower infection rate (30% to 50% of the first waves); (2) a longer incubation period (double to ten times of the first waves); a higher quarantine rate in general; (4) a higher recovery rate; and (5) a lower mortality rate. For a specific resurgence in a country or region, these findings may vary due to their particular circumstances and driving factors.

In conclusion, the second waves in all countries were associated with weaker interventions but stronger deconfinement activities than their first waves. As a result of the relaxed interventions, the second waves had lower infection rates, higher quarantine rates, and longer incubation periods. These result in longer, higher and broader waves of infections in the second waves, also partially attributed to the improved awareness and practices of self-protection and vaccination in some countries. However, their recovery rates and mortality rates were not necessarily lower than the first waves, particularly when relaxed interventions and restrictions on public activities occurred. This may also be related to much higher and longer infections and the resultant overwhelmed healthcare services.

### How do different severity, number and timing of interventions individually and cumulatively affect the trends of the two waves?

To answer this question, we infer the impact of control and relaxation events on the daily cases, and compare the individual and cumulative impacts of the sequentially implemented events in the first and second waves in the five countries. With the different types (control vs. relaxation), number and implementation timing (the dates) of interventions and public activities implemented over two waves in the five countries (as shown in Tables 3-7), Tables 13-17 show the inferred event impact attributes (the top sections) and the individual impact rate (the fitted rate value) of these events implemented in the first and second waves in each country. The parameter subscripts and superscripts in each table represent the corresponding wave number and the event number, respectively. The top section includes the event-sensitive attributes inferred by the iSPEIQRD model, namely the initial protection rate *α*, the initial deconfinement rate *τ*, and the time delay *ν*. The middle part shows the inferred impact *π* of each event (both control and relaxation events) taking place during a wave period. The events affect the protection rates (*α*_1_ and *α*_2_) and the deconfinement rates (*τ*_1_ and *τ*_2_) in a cumulative way per the step functions (see Equations (8)-(9)), which cause fluctuations of the COVID-19 curves. Lastly, the overall impact of both control and relaxation events on cases is inferred at the bottom of the tables.

For an intuitive demonstration of how each event affects cases and how all events cumulatively affect cases, we align the event impact rates with the actual daily new cases in both waves in each country. First, we align each event in the first and second waves with the daily new cases on the same day in each country, as shown in Figure 1. In these figures, the individual impact rate of each event (the time delay factor has already been considered) is shown by the right hand vertical axis. Second, we align the cumulative impact rate of all events with the daily new cases of the first and second waves in Figure 2.

Below, we analyze the findings in terms of two groups of countries. We provide more explanation of how the sequentially implemented control and relaxation events individually and cumulatively affect the daily cases in each wave, why two waves demonstrate different dynamics and trends in each country, how control and relaxation events generate opposite effects, either suppressing or speeding up the COVID-19 curves, and how the inappropriate relaxation such as early relaxation and late intervention may have contributed to the wave differences and COVID-19 resurgence in the two groups of countries with different virus mutations and vaccination conditions.

In **Germany**, first, we discuss the wave differences in terms of the epidemiological attributes and event attributes. As shown in Table 13, a higher proportion of individuals moves directly to the protected compartment (group) in the second wave than in the first one, indicating more people had self-protection consciousness (e.g., staying at home or at other protected status such as immune or quarantined) after experiencing the first wave. This is also consistent with the reduced infection rate shown in Table 8. The two waves have very close and low-level initial deconfinement rates *τ*_1_ and *τ*_2_, reflecting their similar average populations returning from the protected group to the susceptible compartment (e.g., losing immunity and returning to workplace, etc.) at the beginning of the two waves when there were no other relaxation events. In the second wave, there is a longer time delay for the event to take effect. Five control events and three relaxation events are included in the first wave, while three control events and one relaxation event are included in the second wave. In particular, 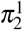, as a large protest without wearing masks and maintaining social distancing, happened at the beginning of the second wave, causing deconfinement and promoting the resurgence. The interventions *almost shutdown* (with impact rate 0.1437) and *stay at home* (0.2056) had the most significant roles in the first and second waves, respectively.

Second, we discuss the individual impact of events occurred in the first and second waves and their relations to the two wave differences. Figure 1 shows the impact of each control or relaxation event on daily cases in the two waves. Aligning the scale of daily new cases and individual event impact in Figure 1 for the first and second waves shows that the events enforced in the first wave had higher containment impact than those implemented in the second wave. The first wave was quickly contained by five powerful control events implemented in the initial stage of the outbreak when the cases were low. In contrast, one significant relaxation event (a protest) was implemented in the early stage of the second outbreak, followed by two strong interventions when the cases were already high, which did not effectively contain the infection quickly, leading to a longer period, a higher peak value, and more cases in the second wave. The result in Figure 1 also explains the significant difference between cases and between event impacts over the two waves shown in Figure 2.

Third, we further discuss the cumulative impact of all different interventions on the wave patterns and differences between two waves. The German government had contradictory response strategies during the two waves, producing different effects on case movements as shown by the protection rate and the deconfinement rate. Figure 2 further shows the respective cumulative effects of all sequentially-implemented events in the two waves on the daily case movement. In the first wave, immediate and strict interventions were implemented, which were mainly adopted in the early stage of the outbreak and generated an immediate effect on flattening the curve. Relaxation measures were then made after the epidemic was substantially contained, and the deconfinement rate *τ*_1_ remained low in the whole first wave. In contrast, in the second wave in Germany shown in Figure 2, interventions lagged behind, probably because of restriction fatigue and concerns about socioeconomic recovery, the effectiveness of these late interventions was also highly discounted. A relaxation event 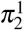 (a protest) took place in the early stage, leading to the increase of the corresponding deconfinement rate *τ*_2_. This caused the exponential increase of daily new cases until the control events 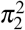 (stay at home), 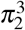 (partial lockdown), and 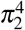 (partial lockdown extension) took effect. Inparticular, the interventions 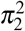 and 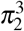 were significant in suppressing the curve rising.

In summary, the different severity, number and timing of sequentially implemented control and relaxation events in the two waves directly contribute to their distinct containment effects on infection in the two waves. The interventions implemented in the two waves led to different epidemic dynamics: the first wave was controlled quickly due to early, strong and more interventions, while the second wave lasted longer and caused a higher peak number and more cases as a result of late, weak and less interventions.

In **France**, first, Table 14 shows the differences between two waves in terms of their intervention impact. The initial protection rate, the deconfinement rate, and the time delay before events taking effect in the second wave were larger than that in the first wave. The increased initial deconfinement rate of the second wave may be due to the early relaxation of social restrictions. As seen in the middle section of the table, five major control events and one relaxation event were implemented in the first wave, in contrast to four major control events and one relaxing event in the second wave. As a result, after a series of interventions enforced at the beginning of the first epidemic, the infection was under control and cases declined. Then the French government reopened businesses 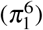) from 12 May. This significant early relaxation may have led to the increase of the initial deconfinement rate in the second wave.Second, we analyze the individual influences of control and relaxation interventions implemented in the two waves on the different wave behaviors and results. The individual effects of control and relaxation events in the two waves in France are shown in Figure 1. In the first wave, five strong control interventions (including school closure 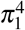 and lockdown 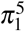) implemented in the very initial stage of the outbreak when the cases were very low generated immediate strong impact on containing cases and curves at the very beginning of the epidemic. Due to the small infectious population in the early stage, the many early and strong interventions quickly contained the first wave. However, in the second wave, the two initial interventions (mask wearing 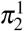 and gathering restriction 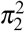) implemented when the cases were already high (much higher than the first wave) were too mild to contain the increase of infections. The cases continuously increased and the outbreak lasted much longer with a much higher peak number of cases. This direction did not change until two strict interventions curfews 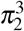 and lockdown 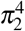 were enforced, which substantially compressed the growth to decline.

Third, we discuss the cumulative impact of all interventions on protection and deconfinement in the two waves. Figure 2 shows the protection rates and the deconfinement rates in both waves. In the first wave, the step-wise early and strong interventions were implemented in the initial stage of the outbreak when the cases were very low. They resulted in a very low deconfinement rate but a much higher protection rate and effectively and quickly contained the first wave. In contrast, the late implementation of fewer strong interventions when the cases were approaching their peak values contributed to a much lower protection rate but a much higher deconfinement rate than the first wave, where deconfinement showed stronger impact than protection. Two initial interventions mask-wearing 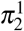 and gathering restrictions 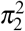 were too soft to influence the case movement, resulted in the higher deconfinement rate than the first wave.

In summary, similar to the scenario in Germany, the different severity, number and timing of control and relaxation interventions in the two waves in France directly contributed to their different containment effects on the two waves, resulting in different case numbers, peak cases and trends in the two waves. Although the two waves were eventually under control, a longer spread period with more infections and higher peak cases was seen in the second wave in comparison to the first. In addition, the overall impact of control interventions in the second wave was much larger than that in the first wave since they took effect only until the curve reached its peak. This explains why a broader and larger population were infected in the longer second wave than the first wave.

In **Italy**, first, Table 15 illustrates the different intervention impacts in two waves. The initial protection rate decreased from 0.0866 in the first wave to 0.0374 in the second wave, while the initial deconfinement rate increased by nearly twice. This is different from Germany and France, which also explains that the first wave was better managed than the second wave in Italy. These inferred event parameters can be further explained by their corresponding interventions. Six control events and three relaxation events were implemented in the first wave, with the wave quickly controlled under these comprehensive strict and decisive interventions. In contrast, the five events implemented in the second wave were only for containment after a big surge of infections, resulting in a softer effect.

Second, Figure 1 explains the individual impact of the events implemented during the two waves on the daily cases. It also explains why the two waves have different numbers of infections and different daily new cases and trends, arising from the different severity, number and timing of control interventions. Similar to Germany and France, strict interventions (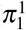 and 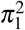) were adopted before the daily new cases reached 1,000. These early and strong interventions implemented in the first wave before the cases rose to a significant level effectively contained the outbreak. In contrast, effective interventions (business shutdown 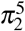) were not taken until the daily new cases exceeded 20,000 in the second wave. The late implementation of four interventions (including school closure 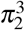) and regional lockdown 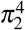) after the cases grew to a substantial level did not take effect. The cases still grew exponentially, which was only compressed after the business shutdown 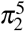 was implemented at avery high case level.

In addition, regarding when to safely reopen businesses, the first wave in Italy shows some hints. From 10 April 2020, the Italian government began to relax the restrictions (cf. Table 5 and Figure 1). Although the daily new cases were still considerable (more than 4,000 on average), the relaxation from 10 April to the end of May in the first wave did not lead to the infection resurgence, showing the possibility of a trade-off between epidemic containment and socioeconomic recovery at the same time after the epidemic has been substantially contained (i.e., in the decline tail stage of an outbreak). Carefully relaxing some restrictions, such as reopening business and relaxing movement restrictions, may not make the infection seriously worse and substantially change the case movement direction as long as (1) such relaxation activities are implemented after an outbreak has been fundamentally controlled and in its decline stage measures; and (2) routine control measures such as self-protection and hygiene (e.g., wearing face masks, hand sanitization, and no hand shaking) and social distancing are still enforced as ‘new normal.’ However, the epidemic may last for a much long period with a long tail, as shown by the case curve in Figure 1).

Third, the cumulative impact of sequential interventions implemented in the two waves on daily new cases further explains the wave differences. Figure 2 shows the protection rates and the deconfinement rates over the two waves. It shows, the first wave was quickly contained since strong interventions but weak deconfinement activities were undertaken at the beginning of the outbreak. As shown in Figure 2, the protection rate *α*_1_ is much higher than the deconfinement rate *τ*_1_. In contrast, in the initial stage of the second wave, the deconfinement rate *τ*_2_ is higher than the protection rate *α*_2_, indicating much relaxed control on the epidemic and leading to an exponential growth of daily new cases. Accordingly, only strict interventions such as full lockdown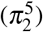 could contain the epidemic, while it would take a longer period and cause more infections since it was implemented too late.

In summary, in Italy, the different behaviors and trends of the two waves were highly associated with their corresponding different severity, number and timing of interventions. Early, strong and more interventions at the initial stage of the outbreak effectively contained the first wave, while late, soft and less interventions resulted a longer and higher resurgence. This case study also shows that balancing epidemic containment and socioeconomic recovery is possible when the epidemic has been substantially contained, which, however, resulted in a longer wave with a long tailed distribution of infections. In addition, the results also show that full lockdown would be mandatory especially when there was an exponential growth of infections and the cases were at a high level.

In **Israel**, Table 16 shows the impact of different interventions in the two waves. Both waves show the similar overall protection rate, while the first wave is associated with a much higher deconfinement rate. This is consistent with some strong interventions such as the strongest intervention synagogue closure 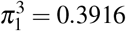, which played a greater role than the strongest intervention night-time curfew 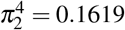 in the second wave. The high deconfinement rate in the first wave is reflected by a highly strong relaxation event easing national lockdown 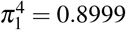, which is much stronger than the highest relaxation easing relaxation 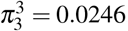. This is also consistent with the very high overall impact of relaxation (0.9047) in the first wave and the very low impact of relaxation (0.024) in the second wave, as shown in Figure 2. The earlier and harsher interventions implemented in the first wave than the second wave better contained the infection of the first outbreak. Consequently, the weaker and later interventions implemented under the context of virus mutations but substantial vaccinations resulted in the higher and denser daily cases.

In **Japan**, the impact of individual events and the cumulative impact of all control and relaxation interventions in both waves show similar patterns and trends to that in Israel, as shown in Table 17. However, the first wave implemented much stronger control interventions in quantity and severity (6 in the first wave vs. 2 in the second wave). Although many control events were conducted in the first wave, the greatest control (state of emergency in six prefectures 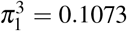) was not strong enough and implemented after the day with peak case number. Two follow-up strong relaxation events(Japanese Naked Festival 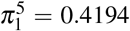 and lifting emergency state in six prefectures 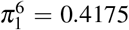) further released the control of infection and triggered the quick resurgence of more infections in the second wave. These are consistent with a relatively low protection rate and a low cumulative impact of control in the first wave, i.e., a high deconfinement rate and a high overall impact of relaxation. In the second wave, as shown in Figure 1, the single control and relaxation events were too weak. The overall protection rate and deconfinement rate are both low, corresponding to very low cumulative impact of control and relaxation, as shown in Figure 2. As a result of weak, late and few control interventions, the second resurgence was associated with high and dense infections, under the context of alpha and delta mutations and partial vaccinations.

### How would more infectious virus mutants influence the second waves under hard or soft interventions and the next 30-day case trends?

As shown in practice, interventions play an important role in changing COVID-19 infection trends and spread particularly when vaccination is low and public health resources are stretched. The first waves in most European countries were successfully contained after rapidly implementing effective interventions. However, in the second waves, most governments hesitated to implement strict control measures after experiencing strict measures in the first wave, resulting in a longer duration and more cases. Here, we simulate two scenarios: whether tightening interventions would change the attributes of the second waves, and what may happen if more infectious virus mutants (such as delta or lambda) appeared in the second wave.

First, we explore the trend of the second waves if the same interventions which successfully contained the first waves were carried forward and compare the simulation results with the actual second waves in each country. This indicates how tightening versus relaxing containment would affect cases. Specifically, we simulate the daily cases by applying the same interventions which were effective in the first wave and observe to what extent the second wave would be controlled with the experience learned from the first wave. The results are shown in Figure 4(a) for the five countries. Table 18 summarizes the change of peak daily cases in the second wave caused by more infectious mutants with strong interventions (i.e.,the same as in the first waves) in these countries.

Second, we simulate the trends of daily cases under more infectious virus mutants in the second wave without changing strategies, i.e., keeping the same relaxation interventions as implemented in the five countries. Figure 4(b) shows the results. Table 19 further shows the change of peak daily cases in the second wave caused by more infectious mutants with soft interventions (i.e., the same as in the second waves) in all countries. In addition to the aforementioned findings, we also find that, when the daily new cases achieve the peak value, the higher the transmissibility, the faster the infection falls. This may be because, with higher transmissibility and looser containment, there would be substantially more cases and many fewer susceptible individuals left, leading to much stronger herd immunity of the population.

Third, we notice that, Israel identified alpha strains in its second resurgence, while the second resurgence of Japan was affiliated with both alpha and delta strains already. Although the waves were incomplete, the estimated patterns of daily cases influenced by virus mutations in Germany, France and Italy (the first three figures in Figure 4(b)) look similar to the original curves of the second waves in Israel and Japan (the two right-bottom diagrams in Figure 1) when alpha and delta appeared already. In Israel, the more infectious virus mutants seem not affect the daily cases (the fifth role in Figure 4) as much as other countries, probably owing to its high vaccination rate and more cautious interventions. The Israeli trends differ from Japan, where the trend of daily new cases would be similar to Germany, France and Italy. This may be related to the weak and late mitigation in Japan, where vaccination didn’t take effect yet, causing high infections.

Lastly, we estimate the trend of the next 30 days after the second waves if more infectious virus mutation happened. In Germany, France and Italy, at the end of December 2020, COVID-19 mutants had been detected in most European countries including these countries. For example, the first mutant was reported on 25 December in Germany, 26 December in France, and 23 December in Italy. We simulate the case movement in the next 30 days following the second wave if virus mutants with different levels of transmission largely spread into communities. Figure 5 shows the estimation results corresponding to three scenarios: mutants with 20%, 50% and 100% infection rate increase, i.e., *β′* = 1.2*β, β′* = 1.5*β*, and *β′* = 2.0*β* for all countries, which simulate more infectious viruses such as delta invade Germany, France and Italy, while Israel and Japan would experience new mutants with a much stronger transmissibility than delta. It shows that all countries including Israel and Japan will be more seriously affected by more infectious virus. Table 20 further shows the change of the estimated daily cases on the 30^*th*^ day after the second wave caused by more infectious mutants in three European countries. Although substantial vaccination and relatively cautious interventions were taken in Israel and the second wave already completed its period, the results do not distinguish it from the three European countries whose second waves were not complete yet. Israel may actually see the highest times of case increase among all countries, this is an interesting finding requiring further exploration. The significant increase by virus mutations may be related the lack of mutant immunity in their community since delta did not appear in the second wave. In contrast, the second wave of Japan was complete and was highly affected by the alpha and delta strains, the more infectious virus than alpha and delta would further increase the infections but as not much as in Israel. This may be owing to the mutation immunity built from the alpha and delta infections in Japan since their second wave already immersed in alpha and delta infections.

### When to safely reopen businesses?

This may be indicated by the overall control-relaxation impact and the protection-deconfinement tradeoff of all interventions and relaxed activities. The protection rate highly exceeds the deconfinement rate over the first waves in Germany, Italy and Israel (Figure 2), resulting in overall control rather than relaxation (0.4732 vs. 0.0399 for Germany, and 0.2908 vs. 0.0003 in Italy) and the possibility of reopening. It is worth noting, at the late stage of the first outbreak in France, Israel and Japan, the deconfinement rate substantially exceeds the protection rate, resulted in resurgence of cases at the end of their first waves. This shows the need of retaining a certain tradeoff between protection and deconfinement over the decline and recovery stage. The estimation of next 30-day case movement under different protection and deconfinement scenarios in Figure 3 further shows it would not be safe to reopen societies at a large scale at the end of the second wave when there would still be considerable infected individuals. With only the limited interventions mixed with relaxation events and insufficient vaccinations, too early or fast relaxation of interventions would definitely return the society to the pandemic outbreak.

### What would be the conditions and costs of living with the virus?

While this research does not directly address this issue, the aforementioned analytical results of relaxing or tightening interventions and activity restrictions at various mutation levels and under unvaccinated and vaccinated conditions provide some insights to this question. To live with the virus: (1) there would be substantial new daily infections even with vaccination, indicating potentially high demand on public health resources including hospitalization and ICU facilities; and (2) strong interventions would still be essential to contain a high number of infections.

### How well does the model predict the future?

We further predict case numbers in the next 10 days following the second wave period, i.e., between 2 Dec. and 11 Dec., 2020 for Germany, France and Italy which were not affected by virus mutations and vaccination; between 28 Mar. and 6 Apr., 2021 for Israel when alpha virus may dominate the infections; and between 21 Jun. and 30 Jun., 2021 for Japan which would be heavily affected by delta. This test verifies the effectiveness of our model in predicting cases with and without virus mutation conditions. Since the cases are highly sensitive to the control and relaxation interventions undertaken, it is not appropriate to make a long-range prediction following the existing conditions. Hence, we make a prediction for the next 10 days only.

The prediction of next 10-day’s active, recovered and deceased cases for the five countries is shown in Figure 15. Two metrics, the root mean square error (RMSE), and mean absolute percentage error (MAPE), evaluate the predictive ability of our model. RMSE in Equation (13) is a standard way to measure the error of a model in predicting quantitative data, and MAPE computes the accuracy as a ratio defined by Equation (14). *y*_*n*_ is the *n*^*th*^-day’s reported value, *y*_−*n*_ is the *n*^*th*^-day’s fitted value by our model, and *N* is the number of observations (in our case *N* = 10). Since active cases, recovered cases and deaths have different orders of magnitude, the generic percentage of MAPE provides a more intuitive view. The RMSE and MAPE values for the daily active cases, the daily recovered cases and the daily deaths in the three countries are shown in Table 21.

**Table 21.**
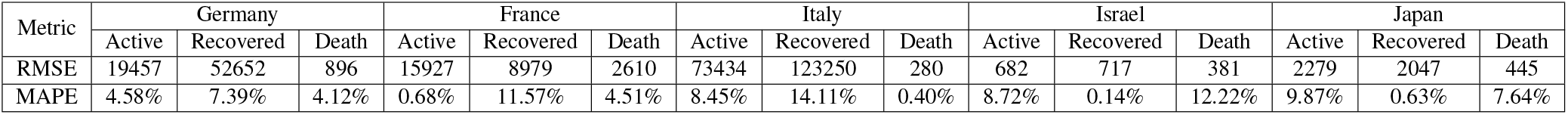
The error of the next 10-day prediction of cases in Germany, France, Italy, Israel and Japan.

| Metric | Germany |  |  | France |  |  | Italy |  |  | Israel |  |  | Japan |  |  |
| --- | --- | --- | --- | --- | --- | --- | --- | --- | --- | --- | --- | --- | --- | --- | --- |
|  | Active | Recovered | Death | Active | Recovered | Death | Active | Recovered | Death | Active | Recovered | Death | Active | Recovered | Death |
| RMSE | 19457 | 52652 | 896 | 15927 | 8979 | 2610 | 73434 | 123250 | 280 | 682 | 717 | 381 | 2279 | 2047 | 445 |
| MAPE | 4.58% | 7.39% | 4.12% | 0.68% | 11.57% | 4.51% | 8.45% | 14.11% | 0.40% | 8.72% | 0.14% | 12.22% | 9.87% | 0.63% | 7.64% |

**Figure 15.**
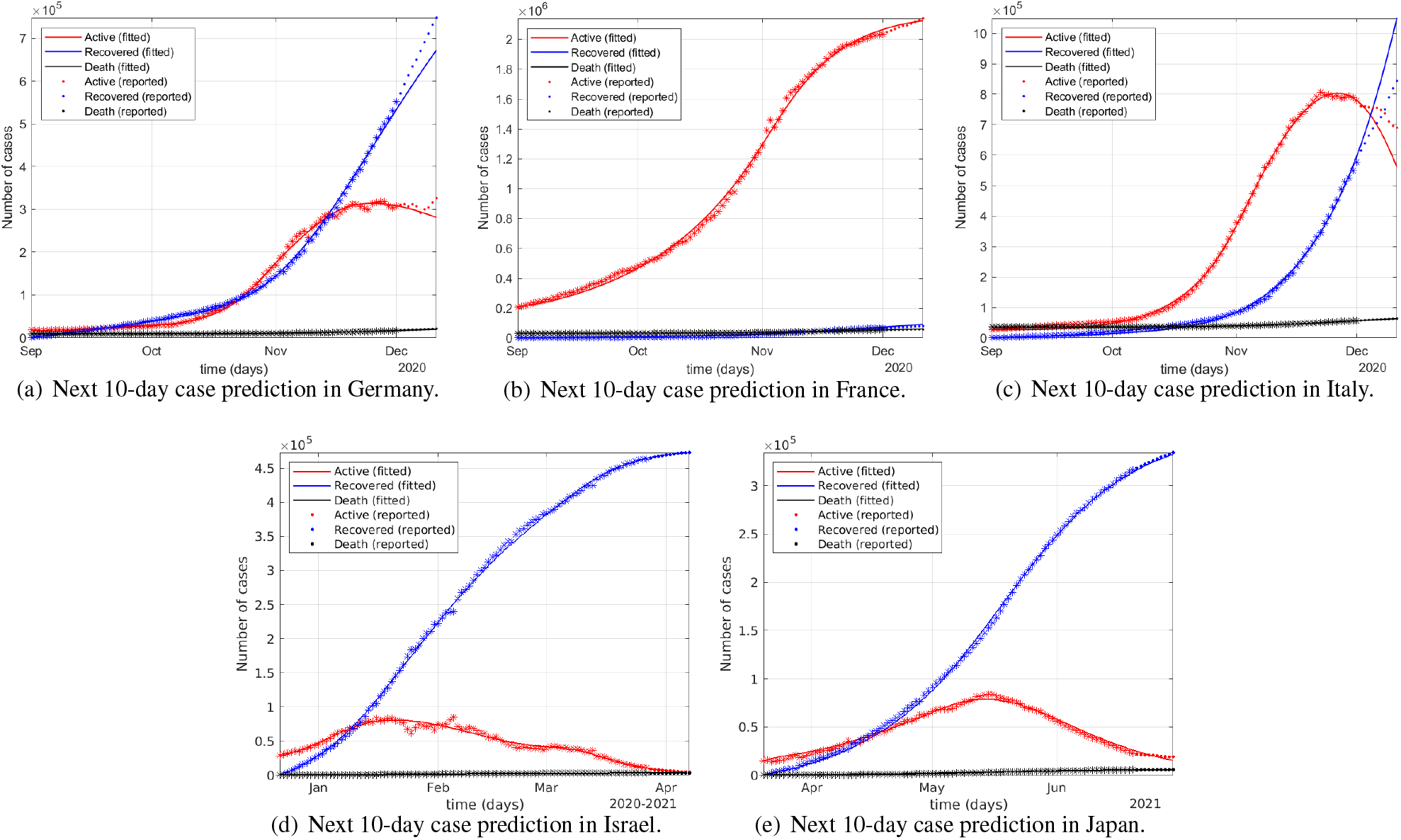
The prediction of the next 10-day active cases, recovered cases and deaths in five countries by the iSPEIQRD model: 2 Dec. - 11 Dec., 2020 for Germany, France and Italy; 28 Mar. - 6 Apr. 2021 for Israel; and 21 Jun. - 30 Jun., 2021 for Japan. The cases during the second wave periods (in star) are the actual, which are used to train the model and infer its parameters. The corresponding lines show the inferred numbers of active, recovered and deceased cases. The actual cases (in dot) of the following 10 days are also shown for comparison.

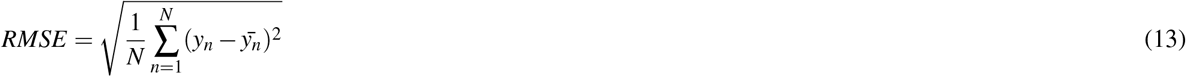

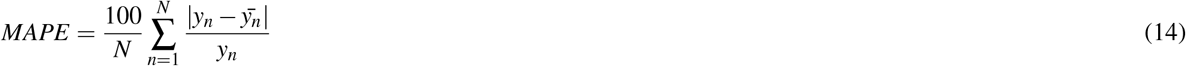

Figure 15 shows the predicted active, recovered and death cases of the next 10 days from our model, precluding the overfitting concern. This result shows the model’s predictability, as evidenced by Table 21, where RMSE presents the average error of each type of cases, and MAPE computes the normalized error about the prediction. The prediction of active and death cases has good performance in all countries as the MAPE values are relatively low (in general, lower than 10% with several less than 1%), which means that our model can generate robust estimates for the case data for the two groups of countries with contrasting scenarios in terms of intervention strategies, virus mutation and vaccination. For the recovered cases, the errors for Germany, France and Italy are relatively higher, with less than 1% for Israel and Japan. This may be due to more statistical noise in the reported recovered case data than in the death case data in the three European countries. As discussed in^4^, the publicly available case data may contain various quality issues such as inconsistency of testing and reporting, under testing and reporting, missing values, and reporting noises, which also significantly challenge prediction.

First, compared to the first waves, the second waves show obviously different epidemiological attributes including more infected cases, and longer incubation periods but lower quarantine rates. These differences result from intervention fatigue, the relaxed and late implementation of interventions, the lagging control measures, and higher deconfinement activities^11^, which lead to more protected individuals rejoining the susceptible population. Second, different levels of interventions have varied effects. Early and strong control interventions push down cases quickly. It is essential to enforce tougher and earlier containment in severe epidemic situations. Third, a too late implementation of interventions or too early reopening likely results in an exponential increase of daily new cases, more serious transmissions, longer incubation and harder, late but longer interventions to get the epidemic under control. Fourth, with more infectious virus mutants, tightening or relaxing interventions would lead to contrasting results of infection, indicating the need for instant implementation of effective interventions at the initial outbreak of a highly infectious mutation. Lastly, different effects and trends of second waves and next 30 days occur under different strategies: maintaining, strengthening or relaxing the interventions; different severity, number and timing of interventions; and under various levels of infectious mutations. We determine the influence of enforcing and relaxing interventions on their wave differences; the better containment of their second waves when more effective interventions were carried forward from their first waves; and the higher risk of infection if more infectious virus mutants are not well controlled.

### Additional findings

Below, we further extend the discussion on the findings and insights of COVID-19 resurgence, including specific wave differences identified in the two groups of countries; the influence of control or relaxation interventions on their wave differences and resurgence; effective containment strategies under different epidemic circumstances and timing; and conditions for business reopening. We address these by comparing the vertical between-wave analyses, horizontal between-country analyses and the first-to-second wave evolution in the three European countries without virus mutations and vaccination with Israel and Japan with virus mutations and vaccination.

First, we summarize the specific wave differences both within and between countries, and also explain how different intervention strategies affect the two waves in these countries which present both similar but also different features. (1) Better contained first waves with different wave patterns: Comparing the first waves in Figure 1, in the early stage, all countries applied many control interventions including lockdowns and night-time curfews which quickly contained the infection spread. However, they had different trends of daily cases, resulting from the different severity, number and timing of the interventions. Germany, Italy, Israel and Japan had a similar long-tailed movement, as a result of their late implementation of many interventions after the outbreak became more severe and the early relaxation of some interventions. In contrast, France implemented and retained several less powerful interventions at the very beginning of the first wave, resulting in a balanced single-peak distribution of cases. (2) Longer and higher second waves: In Figure 1, the Germany’s speedy growth of daily cases was due to their late implementation of two mild interventions, while the four very soft interventions implemented in Italy did not contain the infection until when a strong control measure business shutdown was enforced. In comparison, France had a normal distribution with contained spread within the same period as Germany and Italy by implementing two increasingly stronger control interventions overnight curfew and national lockdown, after two very soft interventions mask wearing and gathering restriction did not control the outbreak. In Israel, the two control interventions implemented when the cases were already high did not contain the infection, and the early easing lockdown further promoted the daily infections. In Japan, only one control measure was implemented at the peak day, which did not contain the spread, resulting in high daily infections. All the second waves show higher peak cases and more daily cases than their first waves across the countries. This may be mainly caused by people’s restriction fatigue and the implementation of inappropriate relaxation interventions in the second waves as well as the more infectious alpha and delta virus strains in Israel and Japan. In addition, the increased test rates in the second waves may also contribute to the higher reported cases. The weak mitigation and the virus mutants may also be associated with the more frequent resurgence of cases in the second wave in Israel and Japan, making it harder to control. (3) Different severity of interventions on first-to-second wave differences: In the first waves, the first control measure was often taken when their daily new cases were under 1,000. In contrast, in the second waves, strong interventions were not adopted until the daily new cases became much higher and approached their peak values (e.g., at around 10,000 in Germany, 5,000 in France and 4,000 in Italy). The impact of similar events differed under the distinct situations of the two waves. For example, lockdown was implemented in both the first and second COVID-19 waves in France. However, the influence on the epidemic was quite different. In the first wave, the inferred lockdown impact rate is 0.0029, which acted more like an enhanced control intervention. In contrast, in the second wave, the impact rate reached 0.4701, which played a decisive role in containing the epidemic. Another example is the *almost shutdown* intervention with an impact rate of 0.2256 in the first wave versus the similar *partial lockdown* with rate of 0.1464 in the second wave in Germany. However, the measure *local lockdown* in the first wave and *regional lockdown* in the second wave in Italy demonstrated similar impact of 0.0307 and 0.0290, respectively. In Israel, the synagogue closure played the most important role in the containment of the first resurgence, while the partial lockdowns did not take more effect than the curfew in the second wave. As partial to full lockdown is often implemented as a preferred (or even first choice) effective intervention in both waves, the effect on containment is sensitive to its severity and timing, full lockdown in the initial stage of an outbreak may maximize the effect of early, short and low-case containment. The previous measures in the second wave in all three countries barely affected the epidemic since they were not effective in terms of severity. (4) Different timing of interventions on first-to-second wave differences: Apart from choosing the suitable severity of an intervention, the timing of enforcing or easing an intervention such as when to reopen business needs to be calculated carefully. Due to relaxed containment, at the end of the second wave, the deconfinement level is higher than that at the end of the first wave in the three European countries, indicating more people rejoined the susceptible group and had higher infection risk. This further shows the importance of managing the frequency and timing (and duration) of interventions. In Israel and Japan, the deconfinement rate at the end of their first wave is much higher than that in the second wave. The relaxations resulted in and explain why the second waves are affiliated with much higher and denser infections.

Second, we discuss general findings on the severity and timing of interventions, the individual and cumulative impact of interventions, and the protection-deconfinement tradeoff of control and relaxation interventions.

- *Intervention severity*: also called strictness or impact level, measures the contribution of each intervention to the containment of an epidemic. Strong and weak interventions result in contrasting results.
- *Intervention timing*: The timing of implementing an intervention differentiates its impact. A strong intervention like full lockdown implemented on the day with peak cases does not quickly flatten the curve as it is implemented in the initial stage of an outbreak.
- *Individual impact*: As shown in Tables 13-15, when a collection of interventions are implemented, each intervention contributes different proportions to influence the overall case movement.
- *Cumulative impact*: While we can measure the overall impacts of control interventions and relaxation interventions, their difference may not distinguish whether an infection or resurgence would be quickly contained.
- Protection-deconfinement tradeoff: The protection and deconfinement rates, shown in both Tables 13-15 and Figure 2, better measure the overall influence of interventions on protection and deconfinement.

Third, we summarize general findings on various scenarios of intervention effects on containment: early vs. late and fast vs. slow containment and discuss conditions for effective containment under different epidemic circumstances and intervention strategies.

- Early containment: An early containment example is shown in the first wave in France in Figure 1.
- Late containment: A late containment example is shown in the second waves in Germany, France, Italy, Israel and Japan in Figure 1.
- *Fast containment*: A fast containment is shown in the second waves in France and Italy and in the first wave in Israel in Figure 1.
- *Slow containment*: A slow containment is shown in the first waves in Germany and Italy and in the second wave in Japan in Figure 1.
- *Effective containment*: For an early and fast containment of an outbreak, called early effective containment, implementing and combining strong interventions such as hotspot-based full lockdown with school closure and stay at home are much more effective than many soft interventions such as mask wearing and social distancing.

Fourth, we summarize the general reasons for wave differences and COVID-19 resurgence and introduce the *protection-deconfinement conflict* to judge whether a resurgence is possible.

- *Wave difference*: As shown in Figure 1, different strictness, timing and number of interventions likely contribute to different epidemic features of different waves in a country and between countries.
- *Resurgence*: In addition to other causes such as imported infections, Figure 1 shows potential causes of resurgence, including early stopping of interventions, premature reopening, and virus mutations.
- *Protection-deconfinement conflict*: Figure 2 shows the protection-deconfinement conflict may indicate the opportunity of resurgence or containment.

Fifth, we discuss the conditions for achieving a tradeoff between containment and relaxation (the containment-relaxation tradeoff) and for business reopening.

- *Containment-relaxation tradeoff in the increase stage*: The second wave in Germany in Figure 1 is an example of imbalance between containment and recovery. “Living with the virus” may be possible in the increase stage of an epidemic only under considerable infection cases with sufficient healthcare resources and moderate socioeconomic activities with good self protection and mild interventions such as mask-wearing, social distancing and large gathering bans).
- *Containment-relaxation tradeoff in the decline stage*: An example of balance is shown in Figure 1.
- *Full reopening*: As shown in Figure 1, fully reopening businesses and societies may be possible only when the cases are extremely low (in comparison with the sufficient affordability of public healthcare resources and the reproduction number is less than 1) and good hygiene and self protection such as social distancing and mask wearing remain valid.

Lastly, we discuss the impact of vaccination by comparing the epidemiological attributes in the three European countries without vaccination and in Israel with substantial vaccination. Japan had a low rate of vaccination close to its late stage of the second wave, which is thus treated similar with Germany, France and Italy.

- *Vaccination impact on infection*: comparing the two waves of Israel in Figure 1, the daily new cases are much more than that in the first wave, although the first and second dose vaccination rates were quite substantial in the second wave. Further research is required to analyze whether vaccination does not substantially prevent infection, i.e., could not reducing the susceptible number substantially but causing more mild or asymptomatic infections.
- *Vaccination impact on more infectious mutation*: The next 30-day estimation under various virus mutations shown in Table 20 does not show the vaccination in Israel could change the infection trends. However, the estimation of Israeli infections sensitive to more infectious virus under soft and strong interventions in Figure 4 shows different patterns from the other four countries with a lower exponential growth.
- *Vaccination impact on the recovery rate*: In Israel, the recovery rate of the second wave is similar to its first wave, indicating their substantial vaccination may not substantially contribute to the recovery.
- *Vaccination impact on the mortality rate*: The mortality rate of the first wave is much higher than that in the second wave. This may indicate the vaccination could significantly contribute to the reduction of deaths.

## Discussion

### Concluding remarks

This research significantly complements the existing work that focuses on either specific epidemiological attributes or the second wave and resurgence without a multi-aspect picture of how infection, virus mutation, interventions, and vaccination interact and affect case resurgence. Our comprehensive comparisons - vertical (between waves), horizontal (across countries), what-if (scenario simulations on second waves), future (30-day trend), contrastive (between vaccinated and unvaccinated), and representative (covering typical ethnic and cultural backgrounds and regions) - in the two representative and critical waves over 2020 and 2021 in Germany, France, Italy, Israel, and Japan show that, in the absence of sufficient vaccination, herd immunity, and effective antiviral pharmaceutical treatments, the widespread early or fast relaxation of interventions including public activity restrictions will result in a COVID-19 resurgence. Importantly, we make the first attempt to simulate the impact of virus mutations (such as delta and lambda) with transmissibility increase of 20 to 100% on the second waves and resurgence infections and the effect of control and relaxation interventions in both vaccinated and unvaccinated countries and both with and without virus mutations (i.e., alpha and delta). We provide deep discussion, comparison, evidence, explanations and insights about important questions and objectives of understanding COVID-19 resurgence, multi-wave evolution and wave differences, the interactions between interventions and epidemic dynamics, the effect of various interventions on resurgences, appropriate intervention strategies on containing virus mutations, and the contrast between vaccinated and unvaccinated communities.

We disclose the relations, behaviors and differences between the first and second waves and how the different epidemiological dynamics were influenced by control and relaxation interventions under different strategies and with more infectious virus mutants. We characterize, simulate and predict the intrinsic epidemiological attributes of the two waves and the impact of enforcing or relaxing interventions (events) on the two-wave cases by a novel intervention-driven compartmental model iSPEIQRD. Our model incorporates both epidemiological and intervention event attributes and their interactions to explicitly explain and estimate the first-to-second wave evolution, the confinement and deconfinement processes, and the future epidemic trend by applying different interventions in the second waves in different European countries. We quantify the influence of control and relaxation events on wave differences with inconsistent case movements. Further, we forecast the possible COVID-19 trends of active, recovered and deceased cases if different control and relaxation interventions were undertaken, to simulate various government responses and control-relaxation intervention combinations. In addition, we evaluate the above aspects by comparing the two groups of countries under different virus mutation and vaccination conditions.

Our analysis characterizes the changes of epidemiological attributes from the first to second wave. More importantly, our experiments also provide a deep understanding of how and why the COVID-19 pandemic unfolds over waves and the wave time periods by analyzing the interactions between the COVID-19 epidemic and significant external factors. We show how various control or relaxation interventions could affect both resurgence and the impact of virus mutations. This study also informs the following understanding and practice of COVID-19 mitigation by interventions. First, the effectiveness of various non-pharmaceutical interventions directly determines how the virus transmits and the effects. It is recommended more effective control measures are always considered in the initial stage of outbreaks to block transmission and control the outbreak as soon and early as possible. When appropriate, priority should be given to strongly effective policies like full lockdowns, business and school shutdowns, strict social distancing and forbidding gathering as informed by the protection and deconfinement rates of the first waves and our simulations. Interventions should not be eased until the virus has been fundamentally contained. Second, the timing of initiating and maintaining interventions is highly deterministic to the spread period and the infection range, further influencing case movements and other aspects. It is much easier and more efficient to control the virus transmission and prevent wide spread and evolution to a regional or national epidemic in the early stage of outbreaks. Third, the strictness of implementing interventions may affect the lifespan of outbreaks. It would be reasonable to say that, if more strict interventions had been implemented at the very beginning of the second wave in the three countries, their resurgences would have been much weaker and many fewer people would have been infected or died.

In addition to the five representative countries evaluated, many other countries have also experienced restriction fatigue and resurgences after managing the first wave of COVID-19. Second and multiple waves have been seen all around the world. Considering the more uncertain and newly emerging factors such as more infectious virus mutants and stranger-to-stranger, household and airborne transmissions, when the mortality rate is still high or unacceptable, a more quantitative and proactive estimation of COVID-19 developments and more timely, effective, targeted and localized restrictions are essential to immediately contain resurgent cases as early as possible and avoid wider and longer regional or national lockdowns and more severe socioeconomic impacts. In balancing epidemic control and societal recovery, policymakers need comprehensive quantitative monitoring and what-if analysis for robust evidence on timely adjusting controls, containing the epidemic, and mitigating societal and psychological impacts.

The findings and analyses in the above sections provide quantitative indication and evidence for governments and policy-makers to systematically manage the severity, number and timing of interventions on resurgences; appropriately balance case development scenarios and intervention strategies; and effectively predict and intervene future COVID-19 resurgence. When the virus is still with unacceptable infection rate and mortality rate, the research also shows that governments and the public have to be prepared for the effects of inappropriate containment and inappropriate timing in managing resurgence and mutations, even under substantially vaccinated conditions. In the stage of virus mutations with high transmissibility and mortality, the research is also useful in the debate on whether elimination (zero COVID, such as the practices undertaken in countries like China), eradication or suppression (“living with the virus” at an acceptable level, like the approaches undertaken in countries like the UK and US and Singapore) and when full reopening would be reasonable in practice.

### Gaps and opportunities

However, our study also has several limitations for further exploration. First, our model assumes an intervention is constant over an entire wave, but it may actually be adjusted over time. For example, the strictness of social distancing, such as restrictions on the number of people participating in indoor events, is often adjusted over the epidemic period according to trends in case development, with harsher restrictions applied in more serious conditions.

Second, in reality, multiple interventions are often enforced simultaneously, interacting with each other and jointly affecting an epidemic’s path. In our modeling, we do not disentangle the impact of multiple events if they were undertaken on the same day since such events are explicitly or implicitly coupled with each other in nature. However, further research could infer the impact of each individual event on case movement for characterization and insight about the positive or negative influence of a specific event.

Third, the time periods of the second waves and resurgence in our case study all end on 1 December 2020 for the three European countries, which is just before the confirmation and spread of more transmissible virus mutants. The epidemiological attributes of the virus mutants and their transmission patterns may significantly differ from their original version, as shown in Israel and Japan. Hence, a question is whether a wave including virus strains such as alpha and delta may introduce significant uncertainty and inconsistency to the modeling and findings in this work^4,48^. In modeling the two waves in Germany, France and Italy, their second waves was not close yet, this wave period selection also leads to the incompleteness of the second waves in our analysis (for example, in Germany and Italy) and may compromise the accuracy of our results, especially for event-related attributes.

Fourth, in simulating the impact of the more infectious virus mutations, we carry forward the epidemiological settings of the original viruses to their mutants. More efforts are required to reflect the epidemiological attributes of specific mutants like delta and lambda. We do not consider the resurgences with omicron since it has been shown much lower mortality, thus the intervention measures applicable to more deadly mutations such as delta and lambda are not applicable. Hence, our studies and findings focus on more critical stages of an infectious disease and its epidemic or pandemic.

Fifth, the level and impact of vaccination combined with interventions on mutated virus spread and external factors such as environments and social activities is not yet quantified in terms of epidemiological attributes and mitigation effect. Further work is required to model whether vaccinated people would have a lower probability of being infected than those unvaccinated. Lastly, the data used may contain various quality issues such as inconsistent, under and missing tests and reporting, small data size, and missing external data such as mobility and environmental factors. For instance, our modeling lacks the degree and extent of actual lockdowns and restrictions and may be biased by the country-based case trends that may significantly vary from the state-based wave behaviors. This could significantly affect modeling performance and results. In addition to enhancing the data quality, modeling low-quality and small COVID case data is a challenging topic for robust forecasting. This also requires future research.

### List of notations

Table 22 lists the notations used in this paper.

**Table 22.** List of notations.

| Notation | Statement |
| --- | --- |
| S | the susceptible |
| P | the protected |
| E | the exposed |
| I | the infected |
| Q | the quarantined |
| R | the recovered |
| D | the death |
| N | the population |
| $\alpha$ | the protection rate |
| $\beta$ | the infection rate |
| $\gamma$ | the incubation rate |
| $\delta$ | the quarantine rate |
| $\lambda$ | the recovery rate |
| $\kappa$ | the mortality rate |
| $\tau$ | the deconfinement rate |
| $\pi_i^j$ | the $j^{th}$ event in the $i^{th}$ wave |
| $\nu$ | the time delay |

## Footnotes

1 Accessed on 30 Nov. 2022 at https://www.worldometers.info/coronavirus.

2 Note, the analysis relies purely on the data-driven discovery without other considerations.

3 We compare the waves across 2020 and 2021, as the waves in 2022 involve significant change of virus nature and effect, and intervention policies.

4 Here, we assume the remaining epidemiological attributes such as mortality rate remain unchanged.

5 The case data sourced from https://github.com/CSSEGISandData/COVID-19/tree/master/csse_covid_19_data

6 https://en.wikipedia.org/wiki/COVID-19_pandemic_in_Germany

7 https://en.wikipedia.org/wiki/COVID-19_pandemic_in_France

8 https://en.wikipedia.org/wiki/COVID-19_pandemic_in_Italy

9 https://en.wikipedia.org/wiki/COVID-19_pandemic_in_Israel

10 https://en.wikipedia.org/wiki/COVID-19_pandemic_in_Japan

11 https://www.who.int/publications/i/item/WHO-2019-nCoV-IHR-Quarantine-2021.1

## References

1. Leung, K., Wu, J. T., Liu, D. & Leung, G. M. First-wave COVID-19 transmissibility and severity in China outside Hubei after control measures, and second-wave scenario planning: a modelling impact assessment. The Lancet (2020).

2. Xu, S. & Li, Y. Beware of the second wave of COVID-19. The Lancet 395, 1321–1322 (2020).

3. Cao, L. & Hou, W. How have global scientists responded to tackling covid-19? medRxiv DOI: 10.1101/2022.08.16.22278871 (2022).

4. Cao, L. & Liu, Q. COVID-19 modeling: A review. medRxiv 1–105, DOI: 10.1101/2022.08.22.22279022 (2022).

5. Cacciapaglia, G., Cot, C. & Sannino, F. Second wave COVID-19 pandemics in Europe: a temporal playbook. Sci. Reports 10, 1–8 (2020).

6. Faranda, D. & Alberti, T. Modeling the second wave of COVID-19 infections in france and italy via a stochastic SEIR model. Chaos: An Interdiscip. J. Nonlinear Sci. 30, 1–25 (2020).

7. Aleta, A. & Moreno, Y. Age differential analysis of COVID-19 second wave in Europe reveals highest incidence among young adults. medRxiv (2020).

8. Grech, V. & Cuschieri, S. COVID-19: A global and continental overview of the second wave and its (relatively) attenuated case fatality ratio. Early Hum. Dev. 105211 (2020).

9. Fan, G. et al. Decreased case fatality rate of COVID-19 in the second wave: A study in 53 countries or regions. Transboundary Emerg. Dis. 68, 213–215 (2021).

10. Lanteri, D., Carco, D., Castorina, P., Ceccarelli, M. & Cacopardo, B. Containment effort reduction and regrowth patterns of the COVID-19 spreading. Infect. Dis. Model. 6, 632–642 (2021).

11. López, L. & Rodó, X. The end of social confinement and COVID-19 re-emergence risk. Nat. Hum. Behav. 4, 746–755 (2020).

12. Mahikul, W., Chotsiri, P., Ploddi, K. & Pan-ngum, W. Evaluating the impact of intervention strategies on the first wave and predicting the second wave of COVID-19 in Thailand: a mathematical modeling study. Biology 10 (2021).

13. Natale, G. D. et al. The evolution of COVID-19 in Italy after the spring of 2020: An unpredicted summer respite followed by a second wave. Int. J. Environ. Res. Public Heal. 17, 1–12 (2020).

14. Zhong, H. & Wang, W. Mathematical analysis for COVID-19 resurgence in the contaminated environment. Math. Biosci. Eng. 17, 6909–6927 (2020).

15. Iftimie, S. et al. First and second waves of coronavirus disease-19: A comparative study in hospitalized patients in Reus, Spain. PLoS ONE 16, 1–13 (2021).

16. Saito, S. et al. First and second COVID-19 waves in Japan: A comparison of disease severity and characteristics: Comparison of the two COVID-19 waves in Japan. The J. Infect. 82, 84–123 (2021).

17. Coccia, M. The impact of first and second wave of the COVID-19 pandemic in society: comparative analysis to support control measures to cope with negative effects of future infectious diseases. Environ. Res. 197, 111099 (2021).

18. Sonabend, R. et al. Non-pharmaceutical interventions, vaccination, and the sars-cov-2 delta variant in england: a mathematical modelling study. The Lancet 398, 1825–1835 (2021).

19. Kodera, S., Rashed, E. A. & Hirata, A. Estimation of real-world vaccination effectiveness of mrna covid-19 vaccines against delta and omicron variants in japan. Vaccines 10, 430 (2022).

20. Aruffo, E. et al. Mathematical modelling of vaccination rollout and npis lifting on covid-19 transmission with voc: a case study in toronto, canada. BMC public health 22, 1–12 (2022).

21. Ge, Y. et al. Untangling the changing impact of non-pharmaceutical interventions and vaccination on european covid-19 trajectories. Nat. Commun. 13, 1–9 (2022).

22. Moore, S., Hill, E. M., Tildesley, M. J., Dyson, L. & Keeling, M. J. Vaccination and non-pharmaceutical interventions for covid-19: a mathematical modelling study. The Lancet Infect. Dis. 21, 793–802 (2021).

23. Ghanbari, B. On forecasting the spread of the COVID-19 in Iran: The second wave. Chaos, Solitons & Fractals 140, 110176 (2020).

24. Renardy, M., Eisenberg, M. & Kirschner, D. Predicting the second wave of COVID-19 in Washtenaw County, MI. J. Theor. Biol. 507, 110461 (2020).

25. Shim, E., Tariq, A. & Chowell, G. Spatial variability in reproduction number and doubling time across two waves of the COVID-19 pandemic in South Korea, February to July, 2020. Int. J. Infect. Dis. 102, 1–9 (2021).

26. Rypdal, K., Bianchi, F. M. & Rypdal, M. Intervention fatigue is the primary cause of strong secondary waves in the COVID-19 pandemic. Int. J. Environ. Res. Public Heal. 17, 1–17 (2020).

27. Huang, B. et al. Integrated vaccination and physical distancing interventions to prevent future COVID-19 waves in Chinese cities. Nat. Hum. Behav. 5, 695–705 (2021).

28. Aleta, A. et al. Modelling the impact of testing, contact tracing and household quarantine on second waves of COVID-19. Nat. Hum. Behav. 4, 964–971 (2020).

29. Ng, V. et al. Projected effects of nonpharmaceutical public health interventions to prevent resurgence of SARS-CoV-2 transmission in Canada. CMAJ 192, E1053–E1064 (2020).

30. Brauner, J. M. et al. Inferring the effectiveness of government interventions against covid-19. Science 371, eabd9338 (2021).

31. Friston, K. J. et al. Second waves, social distancing, and the spread of COVID-19 across the USA. Wellcome Open Res. 5, 1–39 (2021).

32. Schwarzendahl, F. J., Grauer, J., Liebchen, B. & Löwen, H. Mutation induced infection waves in diseases like COVID-19. medRxiv 1–10, DOI: 10.1101/2021.07.06.21260067 (2021).

33. Krueger, T. et al. Risk assessment of covid-19 epidemic resurgence in relation to sars-cov-2 variants and vaccination passes. Commun. medicine 2, 1–14 (2022).

34. Mirri, S., Delnevo, G. & Roccetti, M. Is a COVID-19 second wave possible in Emilia-Romagna (Italy)? forecasting a future outbreak with particulate pollution and machine learning. Computation 8, 1–25 (2020).

35. Fan, G., Song, H., Yip, S., Zhang, T. & He, D. Impact of low vaccine coverage on the resurgence of covid-19 in central and eastern europe. One Heal. 100402 (2022).

36. Gavish, N., Yaari, R., Huppert, A. & Katriel, G. Population-level implications of the israeli booster campaign to curtail covid-19 resurgence. Sci. translational medicine eabn9836 (2022).

37. Lin, L., Zhao, Y., Chen, B. & He, D. Multiple covid-19 waves and vaccination effectiveness in the united states. Int. journal environmental research public health 19, 2282 (2022).

38. Rosenberg, E. S. et al. Covid-19 vaccine effectiveness in new york state. New Engl. J. Medicine 386, 116–127 (2022).

39. Eyre, D. W. et al. Effect of covid-19 vaccination on transmission of alpha and delta variants. New Engl. J. Medicine 386, 744–756 (2022).

40. Pedro, S. A. et al. Conditions for a second wave of COVID-19 due to interactions between disease dynamics and social processes. Front. Phys. 8, 1–9 (2020).

41. Peng, L., Yang, W., Zhang, D., Zhuge, C. & Hong, L. Epidemic analysis of COVID-19 in China by dynamical modeling. arXiv preprint 2002.06563 (2020).

42. Brooks-Pollock, E. et al. The population attributable fraction (PAF) of cases due to gatherings and groups with relevance to COVID-19 mitigation strategies. Phil. Trans. R. Soc. B 376, 1–5 (2020).

43. López, L. & Rodo, X. A modified SEIR model to predict the COVID-19 outbreak in Spain and Italy: simulating control scenarios and multi-scale epidemics. Results Phys. 21, 103746 (2021).

44. Chen, Y.-C., Lu, P.-E., Chang, C.-S. & Liu, T.-H. A time-dependent SIR model for COVID-19 with undetectable infected persons. IEEE Transactions on Netw. Sci. Eng. 7, 3279–3294 (2020).

45. Ambrosio, B. & Aziz-Alaoui, M. On a coupled time-dependent SIR models fitting with New York and New-Jersey states COVID-19 data. Biology 9, 135 (2020).

46. Dong, E., Du, H. & Gardner, L. An interactive web-based dashboard to track COVID-19 in real time. The Lancet Infect. Dis. 20, 533–534 (2020).

47. Hale, T. et al. A global panel database of pandemic policies (Oxford COVID-19 government response tracker). Nat. Hum. Behav. 5, 529–538 (2021).

48. Liu, Q. & Cao, L. Modeling time evolving covid-19 uncertainties with density dependent asymptomatic infections and social reinforcement. Sci. Reports 12, 1–14 (2022).

